# AI-based Hepatic Steatosis Detection and Integrated Hepatic Assessment from Cardiac CT Attenuation Scans Enhances All-cause Mortality Risk Stratification: A Multi-center Study

**DOI:** 10.1101/2025.06.09.25329157

**Authors:** Jirong Yi, Krishna K. Patel, Robert J.H. Miller, Anna M. Marcinkiewicz, Aakash Shanbhag, Waseem Hijazi, Naga Dharmavaram, Mark Lemley, Jianhang Zhou, Wenhao Zhang, Joanna X. Liang, Giselle Ramirez, Valerie Builoff, Leandro Slipczuk, Mark Travin, Erick Alexanderson, Isabel Carvajal-Juarez, René R. S. Packard, Mouaz Al-Mallah, Terrence D. Ruddy, Andrew J. Einstein, Attila Feher, Edward J. Miller, Wanda Acampa, Stacey Knight, Viet T Le, Steve Mason, Vinicius F. Calsavara, Panithaya Chareonthaitawee, Samuel Wopperer, Alan Kwan, Lixia Wang, Daniel S. Berman, Damini Dey, Marcelo Di Carli, Piotr J. Slomka

**Author notes:** **Corresponding Author:** Piotr Slomka, PhD, Cedars-Sinai Medical Center, 6500 Wilshire Boulevard, Los Angeles, California 90048. authors contributed equally as first authors (Jirong Yi, Krishna K. Patel).

## Abstract

**Background:** Hepatic steatosis (HS) is a common cardiometabolic risk factor frequently present but under-diagnosed in patients with suspected or known coronary artery disease. We used artificial intelligence (AI) to automatically quantify hepatic tissue measures for identifying HS from CT attenuation correction (CTAC) scans during myocardial perfusion imaging (MPI) and evaluate their added prognostic value for all-cause mortality prediction.

**Methods:** This study included 27039 consecutive patients [57% male] with MPI scans from nine sites. We used an AI model to segment liver and spleen on low dose CTAC scans and quantify the liver measures, and the difference of liver minus spleen (LmS) measures. HS was defined as mean liver attenuation < 40 Hounsfield units (HU) or LmS attenuation < −10 HU. Additionally, we used seven sites to develop an AI liver risk index (LIRI) for comprehensive hepatic assessment by integrating the hepatic measures and two external sites to validate its improved prognostic value and generalizability for all-cause mortality prediction over HS.

**Findings:** Median (interquartile range [IQR]) age was 67 [58, 75] years and body mass index (BMI) was 29.5 [25.5, 34.7] kg/m^2^, with diabetes in 8950 (33%) patients. The algorithm identified HS in 6579 (24%) patients. During median [IQR] follow-up of 3.58 [1.86, 5.15] years, 4836 (18%) patients died. HS was associated with increased mortality risk overall (adjusted hazard ratio (HR): 1.14 [1.05, 1.24], p=0.0016) and in subpopulations. LIRI provided higher prognostic value than HS after adjustments overall (adjusted HR 1.5 [1.32, 1.69], p<0.0001 vs HR 1.16 [1.02, 1.31], p=0.0204) and in subpopulations.

**Interpretations:** AI-based hepatic measures automatically identify HS from CTAC scans in patients undergoing MPI without additional radiation dose or physician interaction. Integrated liver assessment combining multiple hepatic imaging measures improved risk stratification for all-cause mortality.

**Funding:** National Heart, Lung, and Blood Institute/National Institutes of Health.

**Research in context:** *Evidence before this study:* Existing studies show that fully automated hepatic quantification analysis from chest computed tomography (CT) scans is feasible. While hepatic measures show significant potential for improving risk stratification and patient management, CT attenuation correction (CTAC) scans from patients undergoing myocardial perfusion imaging (MPI) have rarely been utilized for concurrent and automated volumetric hepatic analysis beyond its current utilization for attenuation correction and coronary artery calcium burden assessment. We conducted a literature review on PubMed and Google Scholar on April 1^st^, 2025, using the following keywords: (“liver” OR “hepatic”) AND (“quantification” OR “measure”) AND (“risk stratification” OR “survival analysis” OR “prognosis” OR “prognostic prediction”) AND (“CT” OR “computed tomography”). Previous studies have established approaches for the identification of hepatic steatosis (HS) and its prognostic value in various small-scale cohorts using either invasive biopsy or non-invasive imaging approaches. However, CT-based non-invasive imaging, existing research predominantly focuses on manual region-of-interest (ROI)-based hepatic quantification from selected CT slices or on identifying hepatic steatosis without comprehensive prognostic assessment in large-scale and multi-site cohorts, which hinders the association evaluation of hepatic steatosis for risk stratification in clinical routine with less precise estimates, weak statistical reliability, and limited subgroup analysis to assess bias effects. No existing studies investigated the prognostic value of hepatic steatosis measured in consecutive patients undergoing MPI. These patients usually present with multiple cardiovascular risk factors such as hypertension, dyslipidemia, diabetes and family history of coronary disease. Whether hepatic measures could provide added prognostic value over existing cardiometabolic factors is unknown. Furthermore, despite the diverse hepatic measures on CT in existing literature, integrated AI-based assessment has not been investigated before though it may improve the risk stratification further over HS. Lastly, previous research relied on dedicated CT scans performed for screening purposes. CTAC scans obtained routinely with MPI had never been utilized for automated HS detection and prognostic evaluation, despite being readily available at no additional cost or radiation exposure.

*Added value of this study:* In this multi-center (nine sites) international (three countries) study of 27039 consecutive patients undergoing myocardial perfusion imaging (MPI) with PET or SPECT, we used an innovative artificial intelligence (AI)-based approach for automatically segmenting the entire liver and spleen volumes from low-dose ungated CT attenuation correction (CTAC) scans acquired during MPI, followed by the identification of hepatic steatosis. We evaluated the added prognostic value of several key hepatic metrics—liver measures (mean attenuation, coefficient of variation (CoV), entropy, and standard deviation), and similar measures for the difference of liver minus spleen (LmS)—derived from volumetric quantification of CTAC scans with adjustment for existing clinical and MPI variables. A HS imaging criterion (HSIC: a patient has moderate or severe hepatic steatosis if the mean liver attenuation is < 40 Hounsfield unit (HU) or the difference of liver mean attenuation and spleen mean attenuation is < −10 HU) was used to detect HS. These hepatic metrics were assessed for their ability to predict all-cause mortality in a large-scale and multi-center patient cohort. Additionally, we developed and validated an eXtreme Gradient Boosting decision tree model for integrated liver assessment and risk stratification by combining the hepatic metrics with the demographic variables to derive a liver risk index (LIRI). Our results demonstrated strong associations between the hepatic metrics and all-cause mortality, even after adjustment for clinical variables, myocardial perfusion, and atherosclerosis biomarkers. Our results revealed significant differences in the association of HS with mortality in different sex, age, and race subpopulations. Similar differences were also observed in various chronic disease subpopulations such as obese and diabetic subpopulations. These results highlighted the modifying effects of various patient characteristics, partially accounting for the inconsistent association observed in existing studies. Compared with individual hepatic measures, LIRI showed significant improvement compared to HSIC-based HS in mortality prediction in external testing. All these demonstrate the feasibility of HS detection and integrated liver assessment from cardiac low-dose CT scans from MPI, which is also expected to apply for generic chest CT scans which have coverage of liver and spleen while prior studies used dedicated abdominal CT scans for such purposes.

*Implications of all the available evidence:* Routine point-of-care analysis of hepatic quantification can be seamlessly integrated into all MPI using CTAC scans to noninvasively identify HS at no additional cost or radiation exposure. The automatically derived hepatic metrics enhance risk stratification by providing additional prognostic value beyond existing clinical and imaging factors, and the LIRI enables comprehensive assessment of liver and further improves risk stratification and patient management.

## Introduction

Computed tomography attenuation correction (CTAC) scans are obtained routinely during hybrid myocardial perfusion imaging (MPI) to enhance image quality and diagnostic performance.^1^ Although there has been interest in extracting supplementary anatomic information, such as identifying coronary artery calcium, these scans have not been broadly utilized for other clinical purposes. In the United States, more than 5 million MPI studies are performed annually, many with associated CTACs.^2, 3^ Moreover, in general, over 20 million chest CT exams are performed.^4, 5^ Leveraging these existing data sources could substantially enhance their clinical utility for cardiometabolic and other health-related assessments, with the potential for direct impact on patient care.^2, 3^

Hepatic steatosis (HS) is a prevalent cardiometabolic risk factor, affecting an estimated 32.5 million US adults.^6, 7^ It is strongly associated with insulin resistance and an elevated risk of type II diabetes.^8^ Without proper identification and treatment, up to 20% of individuals with HS can progress to liver fibrosis and cirrhosis, markedly increasing the risk of complications such as portal hypertension, hepatocellular carcinoma, and end-stage liver disease.^6, 8^ While liver biopsy is the diagnostic gold standard, its invasiveness and cost limit regular use.^8^ Only two prior studies utilized AI-based automated hepatic quantification over dedicated abdominal CT imaging to evaluate their association with cardiovascular morbidity or mortality.^8, 9^ However, they were limited either in small population size or single-site cohort.^8, 9^

Non-contrast CT provides an opportunity for opportunistic HS detection in patients undergoing imaging for unrelated indications.^8, 9^ However, CTACs obtained with MPI have never been systematically leveraged for this purpose. The burden of cardiometabolic risk factors including obesity, hypertension, dyslipidemia, and insulin resistance, continues to rise in patients undergoing MPI, reflecting the growing confluence of coronary and metabolic disease.^2, 3^ HS identified on hybrid MPI exams could supply additional prognostic insights over established clinical and imaging markers, allowing for a more comprehensive evaluation of cardiometabolic risk.^9^

We aimed to noninvasively detect HS, quantify HS-related biomarkers from CTAC scans, and determine their added value beyond established clinical and MPI-based risk factors for predicting all-cause mortality (ACM) in a large multicenter population. We also developed and validated an AI-based model for integrated hepatic assessment with modifying demographic factors included, introducing a liver risk index (LIRI) to further improve mortality prediction and risk stratification.

## Methods

### Study population

This retrospective study consisted of 17862 consecutive patients who underwent PET/CT MPI at five sites from 2007 to 2022 and 11305 consecutive patients who underwent SPECT/CT MPI at four sites from 2009 to 2021 participating in the REgistry of Fast Myocardial Perfusion Imaging with NExt generation PET and SPECT (REFINE PET SPECT).^10^ Patients without CTAC scans (N=695), without liver or spleen coverage in CTAC scans (N=753), or with incomplete clinical data (N=680) were excluded. Finally, 27039 patients were included. The study followed the Declaration of Helsinki, with institutional review board (IRB) approval obtained from each participating institution. Cedars-Sinai Medical Center’s IRB in Los Angeles, California, granted overall study approval. Baseline demographic and clinical data were sourced from the registries. The primary outcome was ACM, determined via the National Death Index for sites in the United States and administrative databases for Canadian and Italian sites. The overall study design and the cohort creation flowchart are summarized in Figure 1 and 2.

**Figure 1.**
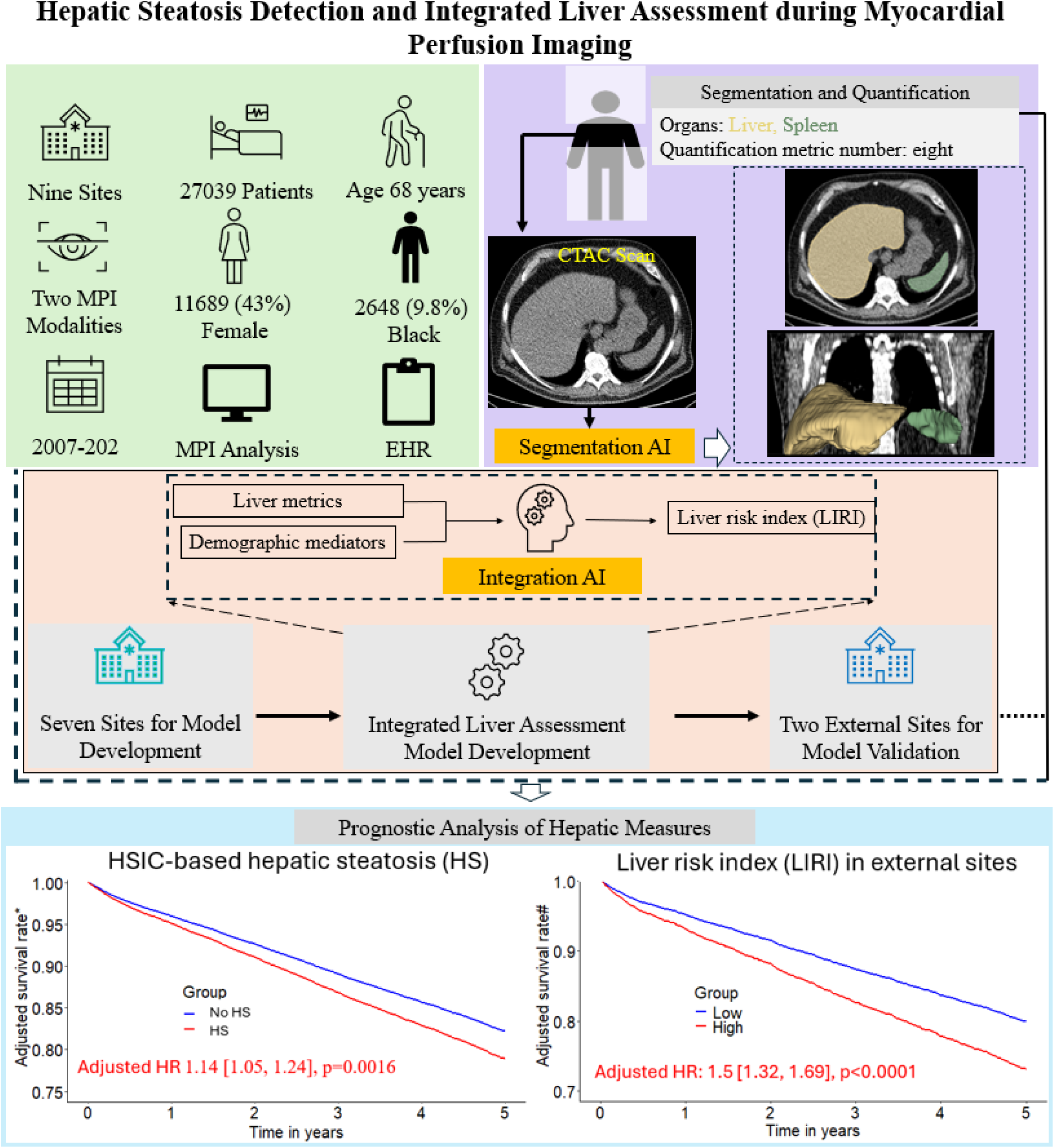
Overview of study design. A previously validated model (TotalSegmentator)^11^ was used to segment liver and spleen for hepatic quantification and hepatic steatosis identification which was subsequently used for risk stratification. Additionally, an eXtreme Gradient Boosting (XGBoost) model was developed and validated for integrated liver assessment, achieving further improvement over individual hepatic measure in risk stratification. * Adjusted for clinical variables, MPI metrics, and all hepatic measures. # Adjusted for clinical and MPI variables. MPI – myocardial perfusion imaging, EHR – electronic health record, CTAC – computed tomography attenuation correction.

**Figure 2.**
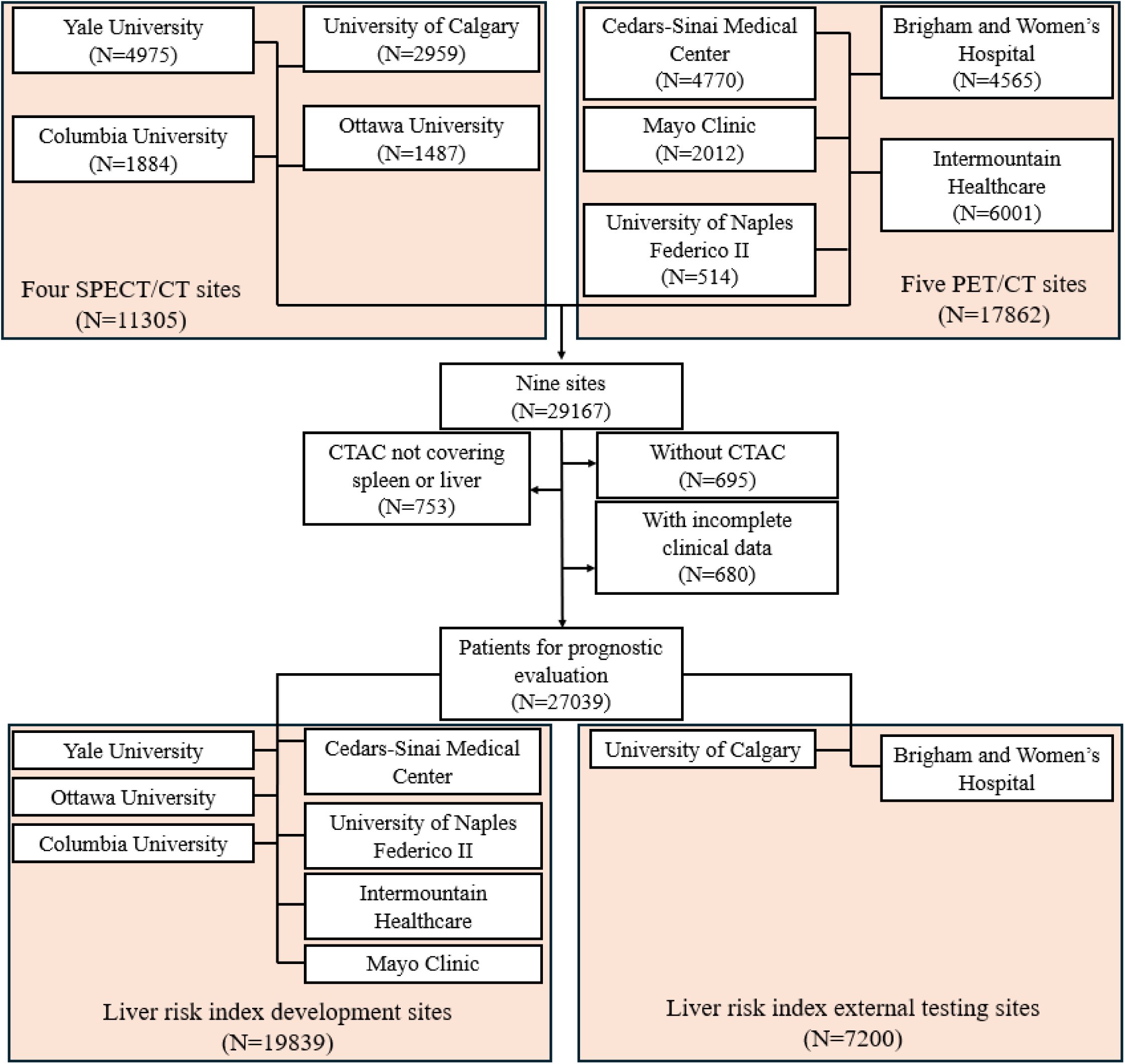
Study cohort creation flowchart with 27039 patients from four SPECT/CT sites and five PET/CT sites. SPECT – single-photon emission computed tomography, CT – computed tomography, PET – positron emission tomography, CTAC – computed tomography for attenuation correction

### CT image acquisition parameters

All CTAC scans were low-dose, non-contrast, and non-electrocardiographically-gated. They were acquired using scanners from different vendors (General Electric, Philips, and Siemens). The slice thickness was 2-5 mm, a tube current of 10-407 mAs and a tube potential of 80-140 kilovoltage peak (kVp). Scans from two sites were conducted with an end-expiratory breath hold while scans from the other seven sites were acquired during normal breathing. An overview of key image acquisition parameters is presented in Supplementary Table 1.

### Hepatic segmentation and quantification

We used a previously validated AI-based CT segmentation model based on nnUnet and TotalSegmentator to automatically segment the liver and spleen volumes from CTAC scans.^11^ The total computation time for segmentation was 81.89 ± 4.54 (mean ± SD) seconds per case. We quantified eight volume-based measures: liver measures (attenuation mean in Hounsfield Units [HU], standard deviation (SD), entropy, and coefficient of variation (CoV)), and differences of liver minus spleen (LmS) measures (attenuation, SD, entropy, and CoV). The CoV was defined as the ratio of SD to mean for measuring the dispersion of tissue density with respect to its mean while the entropy measures the disorder and randomness of the intensity.

### Clinical thresholds

HS diagnosis from CT was defined according to a HS imaging criterion (HSIC): mean liver attenuation < 40 HU, or LmS attenuation < −10 HU as in existing literature.^6, 12^ The maximally selected rank statistic was used to determine cutoffs from the entire population for the new hepatic measures (except HSIC) in prognostic analysis as previously validated cutoffs were not available in the literature.^13^

### Liver risk index for integrated liver assessment

We trained an eXtreme Gradient Boosting (XGBoost) decision tree model for integrated liver assessment by using a supervised binary classification task for all-cause mortality prediction, and combining eight hepatic metrics and demographics (sex, age, and BMI) as input to derive LIRI between zero and one for mortality association evaluation. Model performance was rigorously validated by an external testing scheme. Cases from seven sites (Cedars-Sinai Medical Center, Mayo Clinic, Intermountain Healthcare, University of Naples Federico II, Yale University, Columbia University, and University of Ottawa) were used to develop the XGBoost model, while the remaining sites (University of Calgary, and Brigham and Women’s Hospital) were used for testing (Supplementary Figure 1). Searching space of key hyperparameters for hyperparameter tuning and the final selected hyperparameters were presented (Supplementary Table 2). More details about model hyperparameter tuning/training/external testing are presented in Supplementary Methods. Maximally selected rank test-based cutoff was used for mortality risk stratification with liver risk index (LIRI) and patients with LIRI below the cutoff were used as reference group.^13^

### Clinical data and imaging risk factors

Clinical data at time of imaging was provided from participating centers. Automated quantification of stress total perfusion deficit (TPD) and left ventricular ejection fraction (LVEF) from myocardial perfusion imaging (MPI) scans was performed using specialized software Cedars-Sinai Cardiac Suite at the core lab.^14, 15^ For PET patients, the myocardial flow reserve (MFR) was also quantified. Coronary artery calcium (CAC) score was quantified automatically from CTACs by a previously validated AI-based CT segmentation model.^16^

### Statistical analysis

Continuous variables are reported as median (interquartile range), while categorical variables are expressed as frequency (%). An independent t-test or Wilcoxon rank sum test was applied for comparing continuous variables and Pearson’s chi-squared test or Fisher’s exact test for categorical variables. Kaplan-Meier curves were plotted to evaluate the prognostic value of various hepatic measures and compared using log-rank test. Multivariable Cox proportional hazards models were fitted to assess the association of HSIC-based HS and other hepatic metrics, including LIRI with ACM, with time to all-cause mortality. These models were adjusted for sex, age, body mass index (BMI), hypertension, diabetes mellitus, dyslipidemia, family history of coronary artery disease (CAD), smoking, stress TPD, stress LVEF, CAC score, and imaging modality). Pearson correlation coefficients between hepatic measures and baseline characteristics were calculated, and variance inflation factor (VIF) was used to avoid multicollinearity. These variables were selected *a priori* based on clinical judgement. Abnormal perfusion, abnormal LVEF and coronary microvascular disease were defined as stress TPD ≥ 5%, LVEF < 50%, and myocardial flow reserve (MFR) < 2, respectively. The proportional hazards and linearity assumptions of the Cox model were assessed using Schoenfeld residuals and Martingale residuals plots, respectively. All tests were two-sided and statistical significance was set at p < 0.05. The analyses were conducted using package in RStudio version 4.3.2.

### Role of funding source

The funders of this study had no role in study design, data collection/analysis/interpretation, or writing of the manuscript.

## Results

### Population characteristics

Median age of the overall cohort was 67 [58, 75] years and 15350 (57%) were male (Table 1). Most of the patients (95%) had liver and spleen covered in the CT scans (Figure 2). HSIC-based HS was detected in 6579 (24%) of patients. During a median follow-up of 3.58 [1.86, 5.15] years, there were 4836 (18%) deaths. HSIC-based HS patients had lower liver attenuation (32 [24, 37] vs 51 [47, 56], p<0.0001) and lower LmS attenuation (4 [−5, 15] vs 14 [9, 19], p<0.0001). HSIC-based HS patients were younger (64 [56, 71] vs 68 [59, 76], p<0.0001), and had higher prevalence of diabetes (2921 (44%) vs 6029 (29%), p<0.0001), and obesity (4993 (76%) vs 7896 (39%), p<0.0001) (Table 1).

**Table 1.** Baseline characteristics in the full population and stratified by hepatic steatosis as defined by hepatic steatosis imaging criterion.

| Characteristics | Overall<br>N = 27039 | Without HS<br>N = 20460 | With HS<br>N = 6579 | p-value |
| --- | --- | --- | --- | --- |
| Age [year] | 67 (58, 75) | 68 (59, 76) | 64 (56, 71) | **<0.0001** |
| Age ≥ 60 years | 19333 (72%) | 15096 (74%) | 4237 (64%) | **<0.0001** |
| Sex |  |  |  | **<0.0001** |
| Female | 11689 (43%) | 9287 (45%) | 2402 (37%) |  |
| Male | 15350 (57%) | 11173 (55%) | 4177 (63%) |  |
| Body mass index [kg/m<sup>2</sup>] | 29.5 (25.5, 34.7) | 28.2 (24.8, 32.6) | 34.6 (30.1, 40.5) | **<0.0001** |
| Body mass index ≥ 30 kg/m<sup>2</sup> | 12889 (48%) | 7896 (39%) | 4993 (76%) | **<0.0001** |
| Modality |  |  |  | **<0.0001** |
| Positron emission tomography | 16740 (62%) | 12383 (61%) | 4357 (66%) |  |
| Single photon emission computed tomography | 10299 (38%) | 8077 (39%) | 2222 (34%) |  |
| Liver attenuation [HU] | 48 (40, 54) | 51 (47, 56) | 32 (24, 37) | **<0.0001** |
| Liver SD [HU] | 44 (31, 56) | 42 (30, 53) | 51 (35, 67) | **<0.0001** |
| Liver entropy [bit] | 7.49 (6.97, 7.85) | 7.44 (6.92, 7.77) | 7.70 (7.16, 8.11) | **<0.0001** |
| Liver CoV | 0.89 (0.61, 1.30) | 0.80 (0.56, 1.07) | 1.62 (1.06, 2.39) | **<0.0001** |
| LmS attenuation [HU] | 13 (7, 18) | 14 (9, 19) | 4 (-5, 15) | **<0.0001** |
| LmS SD [HU] | -2.3 (-5.3, 0.0) | -2.2 (-5.0, -0.1) | -2.7 (-6.8, 0.3) | **<0.0001** |
| LmS entropy [bit] | -0.08 (-0.17, 0.00) | -0.08 (-0.17, 0.00) | -0.08 (-0.17, 0.02) | 0.0008 |
| LmS CoV | -0.30 (-0.62, -0.14) | -0.32 (-0.57, -0.18) | -0.16 (-1.23, 0.28) | **<0.0001** |
| Race |  |  |  | **<0.0001** |
| American Indian | 92 (0.3%) | 70 (0.3%) | 22 (0.3%) |  |
| Asian | 580 (2.1%) | 477 (2.3%) | 103 (1.6%) |  |
| Black | 2648 (9.8%) | 2110 (10%) | 538 (8.2%) |  |
| Pacific Islander | 149 (0.6%) | 121 (0.6%) | 28 (0.4%) |  |
| White | 17186 (64%) | 12725 (62%) | 4461 (68%) |  |
| Unknown | 6384 (24%) | 4957 (24%) | 1427 (22%) |  |
| CAC score [AU]# | 2.08 (0.00, 2.95) | 2.16 (0.00, 2.98) | 1.81 (0.00, 2.84) | **<0.0001** |
| Hypertension | 20096 (74%) | 14918 (73%) | 5178 (79%) | **<0.0001** |
| Diabetes mellitus | 8950 (33%) | 6029 (29%) | 2921 (44%) | **<0.0001** |
| Dyslipidemia | 17645 (65%) | 13297 (65%) | 4348 (66%) | 0.1035 |
| Family history of CAD | 8030 (30%) | 6141 (30%) | 1889 (29%) | 0.0444 |
| Smoking | 4644 (17%) | 3542 (17%) | 1102 (17%) | 0.2935 |
| Stress TPD [%] | 4 (2, 10) | 4 (2, 10) | 5 (2, 11) | **<0.0001** |
| Abnormal perfusion (Stress TPD ≥ 5%) | 12341 (46%) | 9155 (45%) | 3186 (48%) | **<0.0001** |
| LVEF [%] | 66 (55, 75) | 66 (55, 75) | 66 (56, 74) | 0.0412 |
| MFR* | 2.26 (1.77, 2.82) | 2.27 (1.77, 2.82) | 2.25 (1.76, 2.84) | 0.6157 |
| Unknown | 10299 | 8077 | 2222 |  |
| MFR < 2* | 6055 (36%) | 4449 (36%) | 1606 (37%) | 0.2709 |
| Unknown | 10299 | 8077 | 2222 |  |
| Death | 4836 (18%) | 3673 (18%) | 1163 (18%) | 0.6131 |
| Death follow up [year] | 3.58 (1.86, 5.15) | 3.55 (1.84, 5.07) | 3.74 (1.96, 5.56) | **<0.0001** |
| Site |  |  |  | **<0.0001** |
| Brigham and Women's Hospital | 4300 (16%) | 3269 (16%) | 1031 (16%) |  |
| University of Calgary | 2900 (11%) | 2383 (12%) | 517 (7.9%) |  |
| Columbia University | 1859 (6.9%) | 1566 (7.7%) | 293 (4.5%) |  |
| Cedars-Sinai Medical Center | 4661 (17%) | 3128 (15%) | 1533 (23%) |  |
| Intermountain Healthcare | 5653 (21%) | 4632 (23%) | 1021 (16%) |  |
| Mayo Clinic | 1942 (7.2%) | 1189 (5.8%) | 753 (11%) |  |
| University of Naples Federico II | 184 (0.7%) | 165 (0.8%) | 19 (0.3%) |  |
| University of Ottawa | 1430 (5.3%) | 1024 (5.0%) | 406 (6.2%) |  |
| Yale University | 4110 (15%) | 3104 (15%) | 1006 (15%) |  |

Bold indicates statistical significance. Continuous variables are reported as median and interquartile range (IQR); categorical variables are reported as number and percentage. \* MFR was only obtained in PET subpopulation. # CAC score was transformed by a logarithmic function $\log_{10}(\text{CAC score}+1)$ .
CI – confidence interval, BMI – body mass index, CAD – coronary artery disease, TPD – total perfusion deficit, LVEF – left ventricular ejection fraction, CAC – coronary artery calcium, CoV – coefficient of variation, LmS CoV – difference of liver CoV minus spleen CoV, LmS SD – difference of liver SD minus spleen SD, LmS entropy – difference of liver entropy minus spleen entropy, MFR – myocardial flow reserve, AU – Agatston unit, HU – Hounsfield unit

HSIC identified higher percentage of HS patients in PET/CT than in SPECT/CT MPI (26% vs 22%, p<0.0001) (Supplementary Table 3). HSIC-based HS was also more likely to be present in male (27% vs 21%, p<0.0001), those with BMI ≥30 kg/m^2^ (39% vs 11%, p<0.0001), those younger than 60 years (30% vs 22%, p<0.0001), diabetic patients (33% vs 20%, p<0.0001), and those with stress TPD ≥ 5% (26% vs 23%, p<0.0001) (Supplementary Table 4). HSIC-based HS prevalence was similar in patients with MFR<2 and those with MFR≥2 (Supplementary Table 4).

### Cutoffs for dichotomization in clinical use

Patients with mean liver attenuation < 40 HU had higher mortality risk, in overall population, and in subgroups of male and female (Figure 3). However, patients with LmS attenuation < −10 HU had lower mortality risk even after adjustment for clinical and MPI variables. The sex-agnostic cutoffs were used in prognostic analysis with Kaplan-Meier curves since the optimal cutoffs from the entire population and different sex subpopulations are similar (Figure 3; Supplementary Table 5). The liver entropy and SD measures had VIF greater than five and thus, were excluded in the Cox regression. The correlation between liver attenuation and BMI was moderately negative, while the other correlations between the other hepatic measures and clinical/MPI variables were weak (Supplementary Figure 2), implying the added information from new measures.

**Figure 3.**
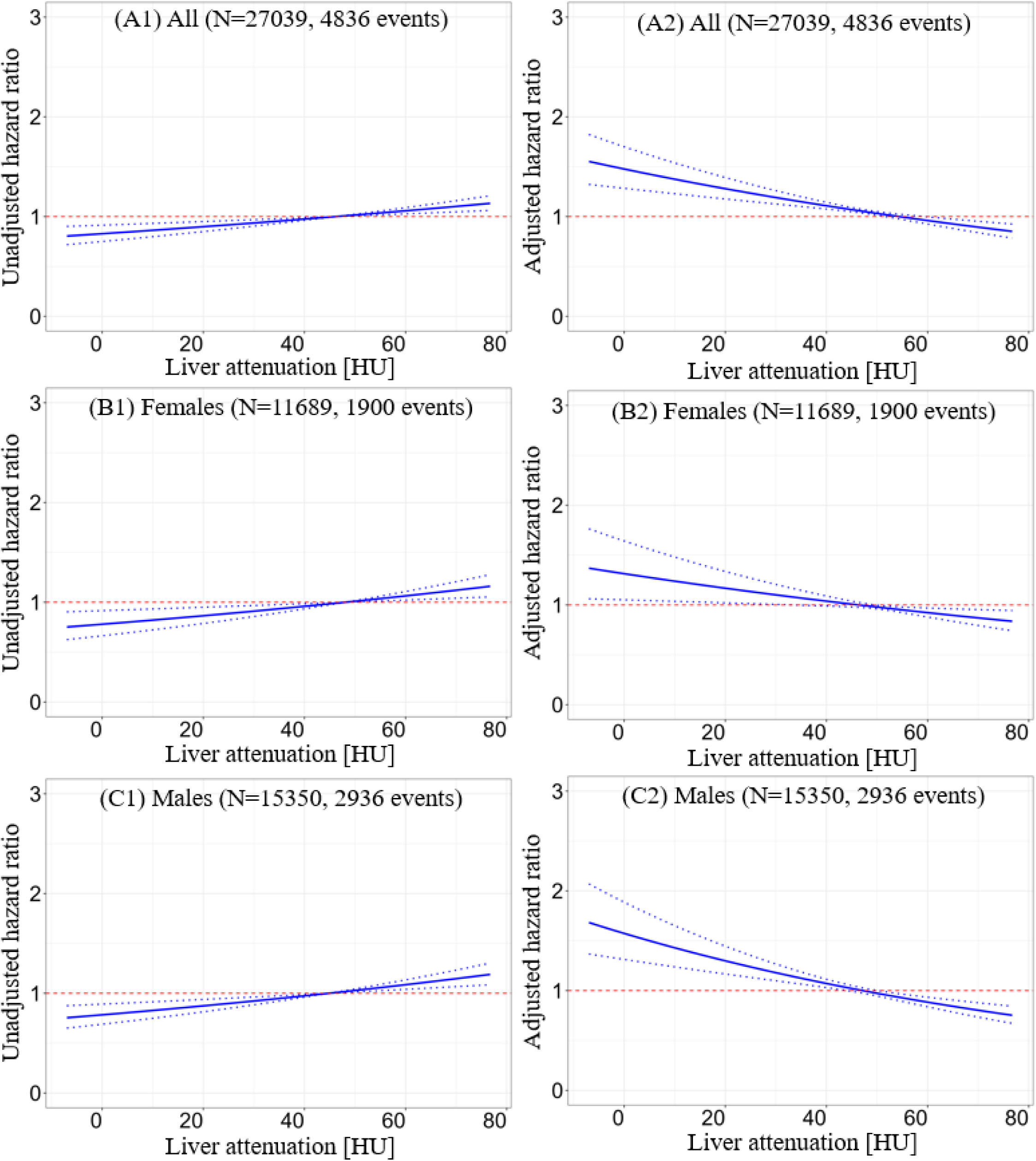
Relation between hazard ratio and hepatic measure. A1, B1, C1: adjusted hazard ratios at different liver minus spleen (LmS) attenuation values with population mean as the reference. A2, B2, C2: adjusted hazard ratios at different liver attenuation values with population mean as the reference. A1-A2: plots from entire population. B1-B2: plots from female subpopulation; C1-C2: plots from male subpopulation. Factors for adjustment included: age, sex, body mass index, diabetes mellitus, dyslipidemia, family history of coronary artery disease, hypotension, smoking, log10(CAC score+1), left ventricular ejection fraction, stress TPD, liver attenuation (or LmS attenuation), liver coefficient of variation (CoV), LmS CoV, LmS entropy, and LmS standard deviation. HU – Hounsfield unit, LmS – liver minus spleen, TPD – total perfusion deficit.

### Association of hepatic measures with all-cause mortality in entire population

HSIC-based HS, low liver CoV, high LmS entropy, and high LmS SD were associated with elevated patient mortality risk (Table 2; Figure 4). After adjustment for clinical, perfusion imaging, and other hepatic risk factors, HSIC-based HS was associated with elevated mortality risk (adjusted HR 1.14 [1.05, 1.24], p=0.0019). Higher liver CoV was associated with decreased mortality risk (adjusted HR 0.84 [0.78, 0.9], p<0.0001) while higher LmS entropy and higher LmS SD were associated with higher mortality rate (adjusted HRs 1.19 [1.06, 1.33] and 1.22 [1.1, 1.35], all p<0.01) (Table 2). Illustrative examples for automatic segmentation, quantification measure and HS identification are shown in Figure 5.

**Figure 4.**
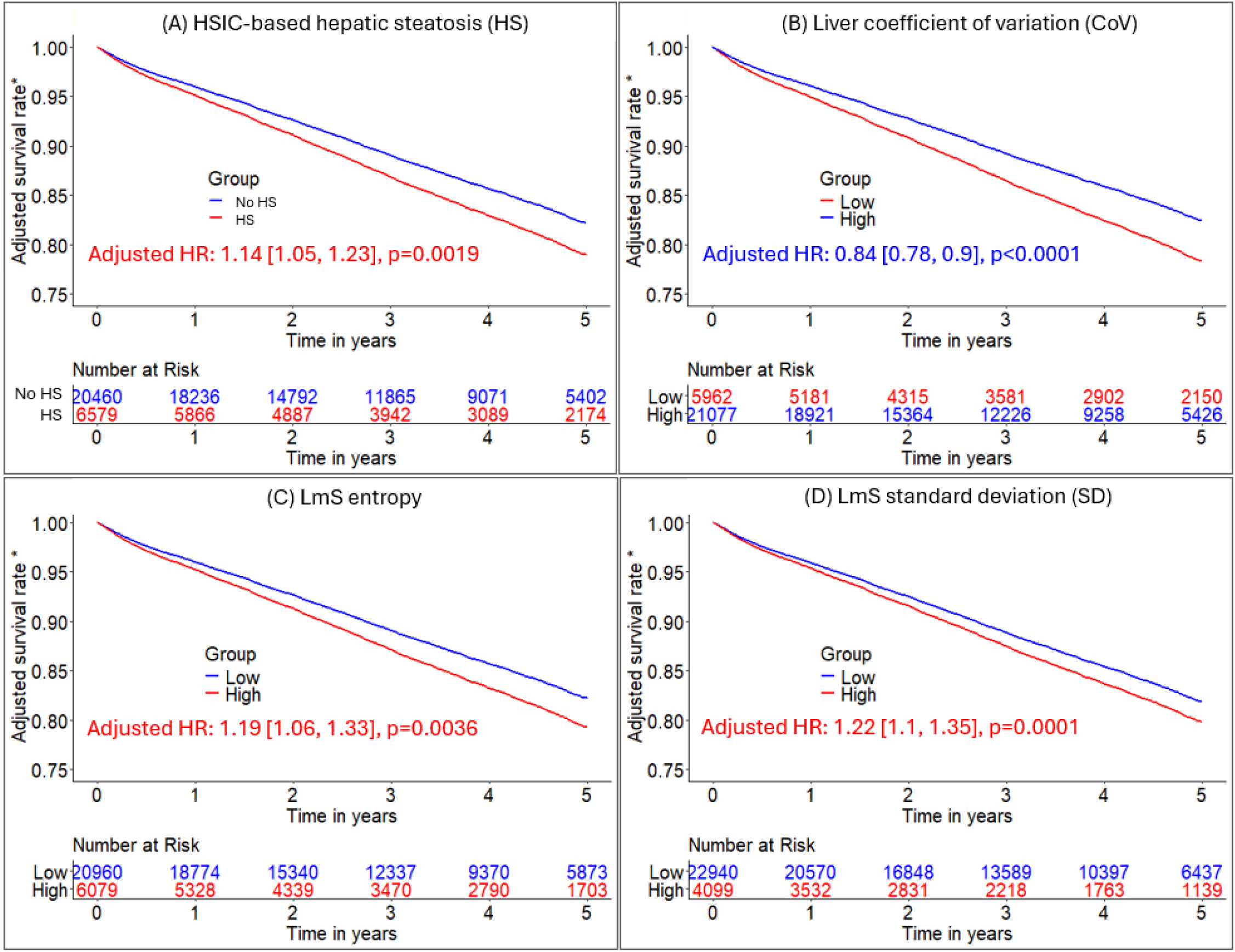
Adjusted Kaplan-Meier curves of hepatic measures for all-cause mortality prognosis. A-D: Kaplan-Meier curves of HSIC-based hepatic steatosis (HS), liver coefficient of variation (CoV), LmS (liver minus spleen) entropy, and LmS CoV for all-cause mortality risk stratification, respectively in MPI cohort of 27039 patients. *Adjusted for clinical/perfusion factors (sex, age, body mass index, hypertension, diabetes mellitus, dyslipidemia, family history of coronary artery disease, smoking, stress total perfusion deficit, left ventricle ejection fraction, coronary artery calcium score, and imaging modality) and all the other hepatic quantifications. Low liver CoV/low LmS entropy/low LmS SD was reference group. Patients without hepatic steatosis measures were in reference group. BMI – body mass index, CAD – coronary artery disease, TPD – total perfusion deficit, LVEF – left ventricular ejection fraction, CAC – coronary artery calcium, LmS – liver minus spleen, MPI – myocardial perfusion imaging, HSIC – hepatic steatosis imaging criterion

**Figure 5.**
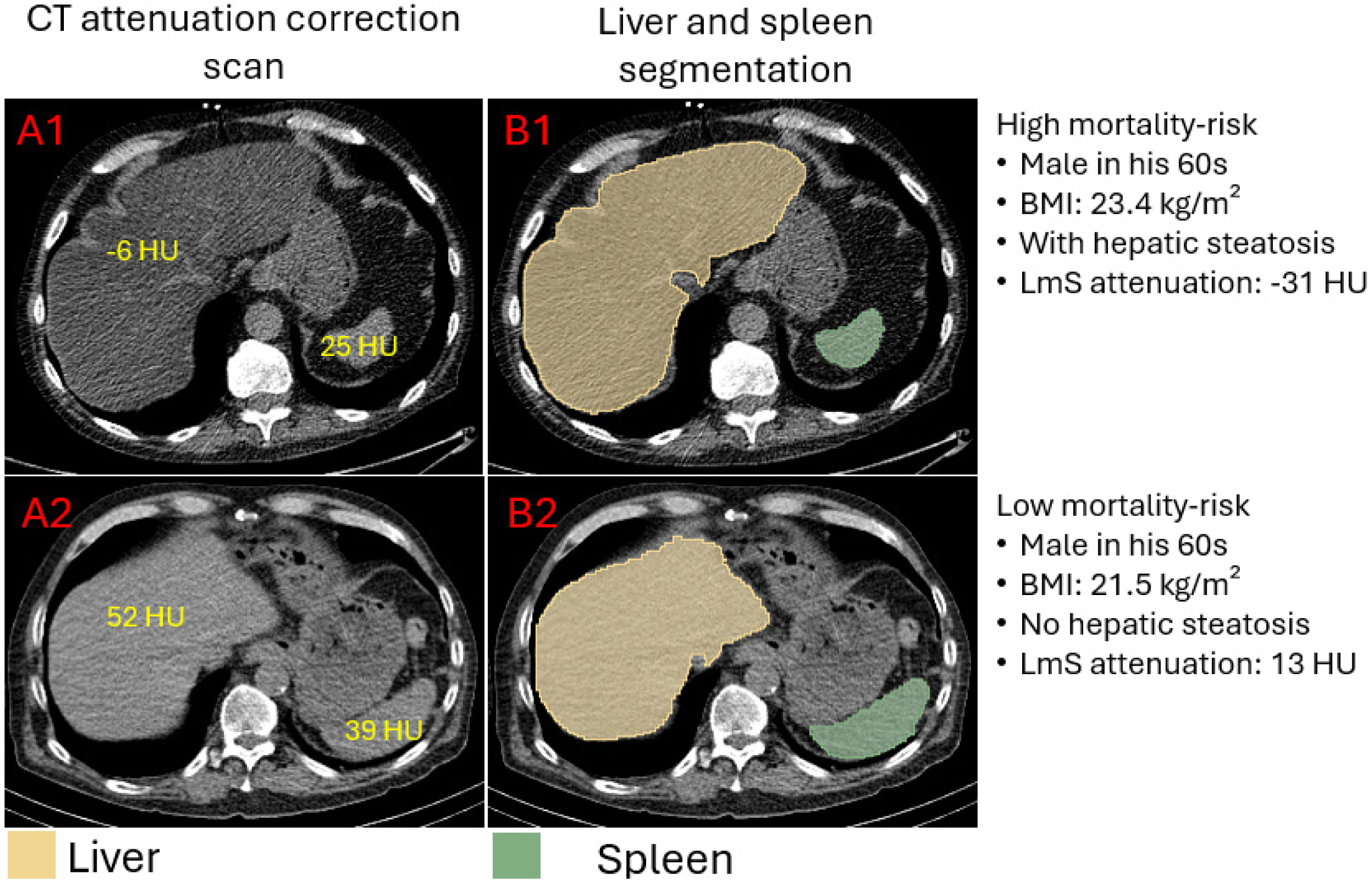
Examples of liver and spleen segmentation from computed tomography attenuation correction (CTAC) scans. A1, B1: axial view slice, and segmentation of liver and spleen from a CTAC scan of a male in his 60s with body mass index 23.4 kg/m^2^, liver attenuation −6 Hounsfield unit (HU), spleen attenuation 25 HU, and difference of liver and spleen (LmS) attenuation −31 HU. The hepatic measure implied the patient had hepatic steatosis (defined as liver attenuation < 40 HU or difference of liver minus spleen attenuation < −10 HU). A2, B2: axial view slice, and segmentation of liver and spleen from a CTAC scan of a male in his 60s with body mass index 21.4 kg/m^2^, liver attenuation 52 Hounsfield unit (HU), spleen attenuation 39 HU, and LmS attenuation 13 HU. BMI – body mass index.

**Table 2.** Unadjusted and adjusted hazard ratios of hepatic measure with categorization in the overall population 27039 patients.

| Factor | Hazard ratio (HR) [95% CI], p value |  |  |
| --- | --- | --- | --- |
|  | Univariable | Multivariable# | Multivariable* |
| Age | **1.05 [1.05, 1.05], <0.0001** | -- | **1.04 [1.04, 1.04], <0.0001** |
| BMI | **0.96 [0.96, 0.97], <0.0001** | -- | **0.98 [0.97, 0.98], <0.0001** |
| Diabetes mellitus | **1.5 [1.42, 1.59], <0.0001** | -- | **1.48 [1.39, 1.57], <0.0001** |
| Dyslipidemia | **1.22 [1.15, 1.3], <0.0001** | -- | **0.8 [0.75, 0.86], <0.0001** |
| Family history | **0.73 [0.68, 0.78], <0.0001** | -- | **0.81 [0.76, 0.87], <0.0001** |
| Sex | **1.29 [1.21, 1.36], <0.0001** | -- | 0.94 [0.88, 1], 0.0624 |
| Hypertension | **1.85 [1.71, 2.01], <0.0001** | -- | **1.24 [1.14, 1.35], <0.0001** |
| log10(CAC score+1) | **1.48 [1.44, 1.51], <0.0001** | -- | **1.18 [1.15, 1.22], <0.0001** |
| LVEF | **2.75 [2.59, 2.91], <0.0001** | -- | **2.03 [1.9, 2.17], <0.0001** |
| Modality | **2.22 [2.04, 2.42], <0.0001** | -- | **1.88 [1.71, 2.05], <0.0001** |
| Smoking | **1.11 [1.03, 1.2], 0.0065** | -- | **1.36 [1.26, 1.47], <0.0001** |
| Stress TPD | **1.99 [1.88, 2.11], <0.0001** | -- | **1.26 [1.18, 1.34], <0.0001** |
| HSIC-based hepatic steatosis | **0.89 [0.84, 0.95], 0.0008** | **1.1 [1.03, 1.19], 0.0067** | **1.14 [1.05, 1.23], 0.0019** |
| High liver CoV | **0.78 [0.73, 0.83], <0.0001** | **0.9 [0.84, 0.96], 0.0013** | **0.84 [0.78, 0.9], <0.0001** |
| High LmS CoV | **0.85 [0.77, 0.93], 0.0008** | **1.18 [1.07, 1.3], 0.001** | 1.06 [0.95, 1.19], 0.2646 |
| High LmS entropy | **1.27 [1.18, 1.37], <0.0001** | **1.4 [1.3, 1.51], <0.0001** | **1.19 [1.06, 1.33], 0.0036** |
| High LmS SD | **1.24 [1.17, 1.33], <0.0001** | **1.34 [1.25, 1.43], <0.0001** | **1.22 [1.1, 1.35], 0.0001** |

Bold indicates statistical significance.

### Association of hepatic measures with all-cause mortality in sub-populations

HSIC-based HS was associated with increased mortality in male patients (adjusted HR 1.21 [1.1, 1.34], p=0.0001), those with age ≥ 60 years (adjusted HR 1.18 [1.08, 1.28], p=0.0003), non-obese patients (adjusted HR 1.21 [1.07, 1.36], p=0.0015), White patients (adjusted HR 1.21 [1.11, 1.33], p<0.0001), non-diabetic patients (adjusted HR 1.31 [1.17, 1.46], p<0.0001), those with stress TPD ≥ 5% (adjusted HR 1.15 [1.04, 1.28], p=0.0062) but not in female, those with age < 60 years, obese patients, Black patients, diabetic patients, and those with stress TPD < 5% (Supplementary Table 6). Higher liver CoV was persistently associated with reduced mortality risk in both patients with MFR < 2 (adjusted HR 0.87 [0.79, 0.95], p=0.0028) and those with MFR ≥ 2 (adjusted HR 0.82 [0.73, 0.91], p=0.0004) (Supplementary Table 6). The Kaplan-Meier curves of HSIC-based HS for mortality risk stratification in different subpopulations are shown in Supplementary Figure 3.

### Integrated liver risk assessment via LIRI and its improved prognostic value

LIRI achieved higher Harrell’s concordance index than HSIC-based HS in external testing data (0.65 [0.64, 0.67] vs 0.45 [0.42, 0.48], p<0.0001). Higher LIRI was associated with higher mortality risk (adjusted HR 1.5 [1.32, 1.69], p<0.0001). LIRI gives larger effective size and better statistical reliability for risk stratification than HSIC-based HS in overall (adjusted HRs 1.5 [1.32, 1.69], p=0.0001 vs 1.16 [1.02, 1.31], p=0.0204; p<0.0001 for likelihood ratio test of corresponding Cox models) and in subgroups (male patients, adjusted HR 1.53 [1.31, 1.79], p<0.0001 vs 1.14 [0.98, 1.34], p=0.0937; non-obese patients, adjusted HR 1.36 [1.16, 1.59], p<0.0001 vs 1.23 [1, 1.51], p=0.0485; all p<0.0001 for likelihood ratio test of Cox models) (Table 3; Figure 6). Low-risk group stratified by LIRI had higher liver attenuation (52 [42, 59] vs 50 [43, 57], p<0.0001) and higher liver CoV (0.61 [0.47, 0.89] vs 0.59 [0.48, 0.82], p=0.0097) (Supplementary Table 7). Liver attenuation and LmS SD contributed most to LIRI (Supplementary Figure 4).

**Figure 6.**
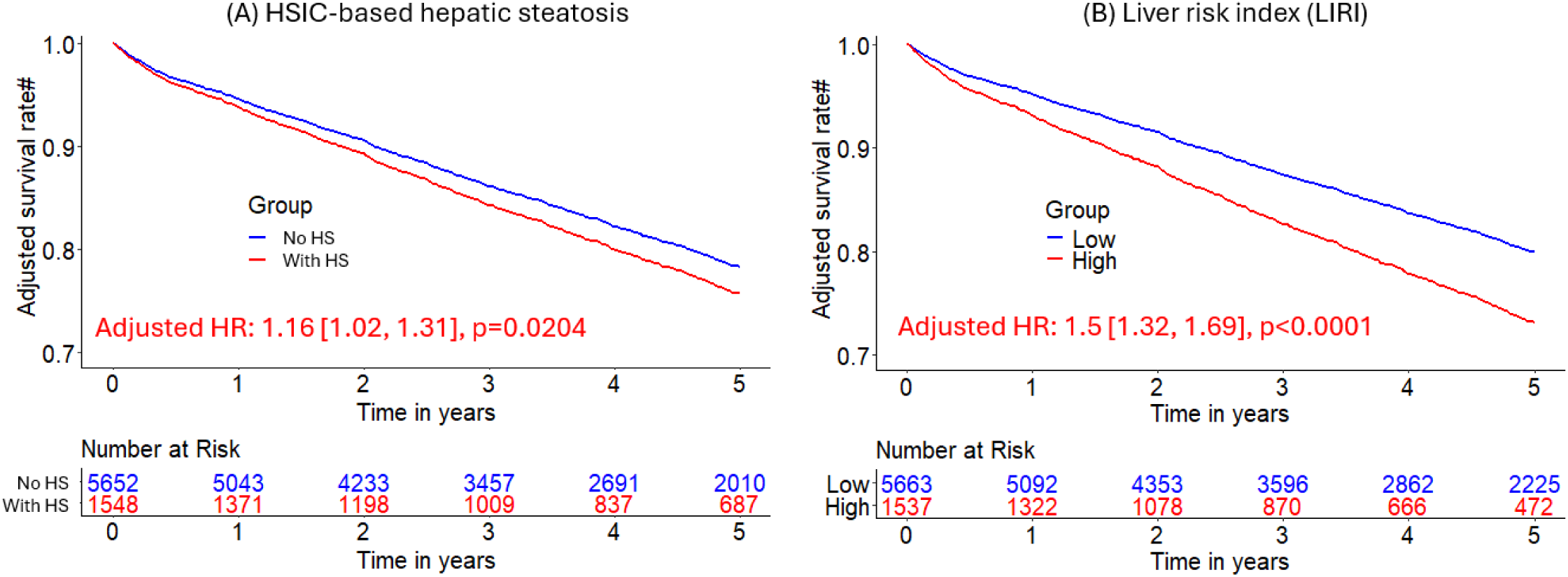
Kaplan-Meier curves of hepatic steatosis imaging criterion (HSIC)-based hepatic steatosis and liver risk index (LIRI) for risk stratification of all-cause mortality in external sites. HS was defined as liver attenuation < 40 HU or difference of liver minus spleen attenuation < −10 HU) in the external sites. # Adjusted for clinical and perfusion variables. Liver risk index (LIRI) was produced by an eXtreme Gradient Boosting model.

**Table 3.** Association evaluation with mortality by HSIC and liver risk index (LIRI). Integrated liver assessment model was developed using cases from 7 sites and tested in 2 remaining sites

| Population |  | Hazard ratio (HR) [95% CI], p value # |  | Likelihood ratio test |
| --- | --- | --- | --- | --- |
|  |  | HSIC | LIRI | p-value |
| Entire (N=7200) |  | **1.16 [1.02, 1.31], 0.0204** | **1.5 [1.32, 1.69], <0.0001** | **<0.0001** |
| Modality | PET site (N=4300) | **1.17 [1.03, 1.33], 0.0148** | **1.46 [1.28, 1.67], <0.0001** | **<0.0001** |
|  | SPECT site (N=2900) | 0.81 [0.54, 1.23], 0.3222 | **1.5 [1.07, 2.08], 0.0174** | **<0.0001** |
| Sex | Female (N=3295) | 1.18 [0.97, 1.44], 0.1057 | **1.46 [1.2, 1.77], 0.0002** | **<0.0001** |
|  | Male (N=3905) | 1.14 [0.98, 1.34], 0.0937 | **1.53 [1.31, 1.79], <0.0001** | **<0.0001** |
| Age | < 60 years (N=2112) | 0.98 [0.74, 1.29], 0.8649 | **1.33 [0.97, 1.82], 0.0814** | **<0.0001** |
|  | ≥60 years (N=5088) | **1.22 [1.07, 1.4], 0.0040** | **1.46 [1.27, 1.67], <0.0001** | **<0.0001** |
| Obesity | Non-obese (N=4019) | **1.23 [1, 1.51], 0.0485** | **1.36 [1.16, 1.59], 0.0002** | **<0.0001** |
|  | Obese (N=3181) | 1.03 [0.88, 1.21], 0.7141 | **1.59 [1.31, 1.94], <0.0001** | **<0.0001** |
| Diabetes | Non-diabetic (N=4861) | **1.28 [1.07, 1.51], 0.0053** | **1.42 [1.21, 1.67], <0.0001** | **<0.0001** |
|  | Diabetic (N=2339) | 1.02 [0.85, 1.22], 0.8336 | **1.5 [1.23, 1.83], 0.0001** | **<0.0001** |
| Race | Black (N=599) | 1.13 [0.79, 1.61], 0.4944 | **1.71 [1.17, 2.49], 0.0051** | **<0.0001** |
|  | White (N=3123) | **1.21 [1.04, 1.4], 0.0129** | **1.41 [1.21, 1.66], <0.0001** | **<0.0001** |
| Stress TPD | < 5% (N=3145) | 1.13 [0.79, 1.61], 0.4944 | **1.33 [1.05, 1.68], 0.0179** | **<0.0001** |
|  | ≥ 5% (N=4055) | **1.21 [1.04, 1.4], 0.0129** | **1.58 [1.36, 1.82], <0.0001** | **<0.0001** |

Bold indicates statistical significance. One external test site did not have race information available in the registry.
Non-obese was defined as BMI less than 30 kg/m<sup>2</sup>, obese was defined as BMI no less than 30 kg/m<sup>2</sup>, abnormal perfusion was defined as stress TPD ≥ 5%, normal perfusion was defined as stress TPD < 5%. #Adjusted for clinical and perfusion factors: sex, age, body mass index, hypertension, diabetes mellitus, dyslipidemia, family history of coronary artery disease, smoking, stress total perfusion deficit, left ventricle ejection fraction, and deep-learning-coronary artery calcium score. Myocardial flow reserve was also included for adjustment at PET. HSIC – hepatic steatosis imaging criterion, BMI – body mass index, TPD – total perfusion deficit

## Discussion

The current study demonstrates the significant prognostic value of AI-based hepatic steatosis detection and integrated hepatic assessment for mortality risk stratification using standard CTAC scans acquired during MPI (both PET and SPECT) in a large international multicenter cohort. Our findings indicate approximately one in four patients undergoing MPI exhibit previously undiagnosed moderate to severe HS, and this subset carries a significantly increased mortality risk, independent of traditional cardiometabolic and imaging risk factors. We also developed and validated an AI-based model for integrated liver assessment by combining the multiple hepatic measures with the demographic variables to derive the LIRI which showed stronger risk-stratification ability than HS overall and in subpopulations. The automated approach leverages routine cardiac imaging protocols, thus providing a method for opportunistic screening of HS without additional radiation exposure or physician effort.

Our study aligns with a growing body of literature recognizing the systemic impact of metabolic dysfunction-associated steatotic liver disease (MASLD), of which HS is the most common manifestation.^8, 17^ The high prevalence of HS (24%) in our cohort of patients referred for MPI underscores the interconnectedness of cardiovascular and metabolic health. Identification of a frequently under-diagnosed HS through existing imaging data presents a significant opportunity for enhanced risk stratification and potentially targeted interventions.^18^ The only two prior studies which utilized AI-based hepatic quantification over dedicated abdominal CT imaging to evaluate the association of HS with increased cardiovascular morbidity or mortality are limited either in small cohort size or single-site cohort.^8, 9^ Our results showed the feasibility and effectiveness of HS screening using AI-derived hepatic parameters from existing cardiac CTAC scans in a multi-center and multi-modality consecutive cohort with the largest scale to date. The observation that HS is independently associated with increased mortality, even in the presence of other major cardiometabolic risk factors, highlights its important role as a prognostic marker in this patient population. This finding suggests that the presence of abnormal CT-based hepatic measures may contribute to all-cause mortality through various mechanisms, including systemic inflammation, dyslipidemia, and insulin resistance.^8, 9^

The use of AI for automated quantification of hepatic measures from CTAC scans represents a significant advancement.^8, 9^ Traditionally, assessment of liver fat often requires scans like ultrasound with controlled attenuation parameters or abdominal CT or dedicated MRI sequences.^17, 19^ Routine manual quantification of liver fat on CT is hindered by the need of standardized measurement protocols, inaccurate delineation for regions of interest and high interrater variability.^8, 9^ Although there has been growing interest in AI-assisted identification of HS using various imaging modalities,^8, 9^ analyses of MPI CTAC scans remain limited despite the fact that these scans are already available at no added cost.^19, 20^ Our approach utilizes data acquired for attenuation correction, incurring no additional radiation dose or workflow disruption. This opportunistic screening strategy has the potential to identify many at-risk individuals who might otherwise remain undiagnosed. Some of the specific hepatic measures we employed, including attenuation, CoV, and entropy, have been previously shown to correlate with the degree of hepatic fat infiltration.^9, 21^

This study provides simple but effective pathways for detecting HS or ruling out HS using CT attenuation maps. Obesity was strongly associated with HS with high positive predictive value (Supplementary Table 8), suggesting potential for identifying individuals with higher HS risk.^22^ On the other hand, diabetes showed high negative predictive value (Supplementary Table 8), consistent with previous studies and useful for ruling out HS when abscent.^22^ HSIC-based HS showed protective effect in unadjusted scenario (Table 2), which could be due to the strong correlation between BMI and HS and reflective of the obesity paradox.^23^ It could also be due to the existence of advanced liver disease such as hepatocellular carcinoma.^24^ These observations further validate the utility of these CT-derived metrics for HS identification and risk stratification. The different associations of HS with mortality risk in different subpopulations highlighted the complexity of using hepatic steatosis for prognostic purpose and the modifying effects of existing factors. These advocated the necessity of dedicated studies of specific populations and motivated us to develop the more robust and unbiased LIRI measure from low dose CT for overall liver health assessment and mortality risk stratification.

The development and validation of the LIRI represent a key contribution of this study. Integrating multiple hepatic CT imaging measures with demographic variables provides a more comprehensive assessment of liver health and demonstrates superior prognostic value compared to HS defined by only attenuation measure. Despite the large number of studies on individual hepatic measures, no prior research has investigated a comprehensive liver assessment that integrates multiple hepatic parameters.^9, 21^ Besides, HS has been shown to have opposite associations with ACM in various cohorts event after adjustment, implying the necessity of integrated liver assessment.^24, 25^ The robust performance of the LIRI in external validation sites further strengthens its potential for clinical translation.

From a clinical perspective, integrating automated hepatic assessment into routine cardiac imaging practices may transform MPI from a purely cardiovascular-focused procedure into a broader cardiometabolic risk evaluation tool.^17^ By identifying previously undiagnosed HS in a substantial proportion of patients, clinicians can adopt a more comprehensive and multidisciplinary management strategy. Early detection could prompt timely interventions, including lifestyle modification, dietary counseling, weight loss, intensified lipid management with statins, and targeted pharmacological therapy to prevent progression to liver fibrosis or hepatocellular carcinoma.^26, 27^ This proactive approach may not only halt or reverse hepatic steatosis progression but also reduce associated cardiometabolic risk and improve patient outcomes.^17^ Additionally, stratifying patients using the LIRI could help identify those at greatest risk, allowing for individualized follow-up protocols and more aggressive preventive measures. Given the rising global prevalence of metabolic syndromes and their associated cardiovascular risks, opportunistic screening approaches offer significant advantages, including early detection, tailored clinical interventions, and ultimately improved cardiovascular outcomes and patient survival.^17^

The large sample size, multi-center nature, and robust external validation of the derived hepatic risk measures are key strengths of our study. Despite this, several limitations need to be considered. First, the diagnosis of HS was based on CT attenuation thresholds (40 HU for liver attenuation and −10 for liver minus spleen attenuation), which, while widely used, are not a direct measure of hepatic fat content like liver biopsy or MRI-based techniques.^17^ The biopsy-based gold standard approach is not widely used in clinical routine due to its invasive nature and high cost.^17^ However, these thresholds have been shown to have good diagnostic accuracy for moderate to severe steatosis and strong relation with hepatic fat content.^6, 12^ When cutoff 40 HU was independently used for risk stratification, higher liver attenuation was associated with increased mortality, consistent with existing studies.^6, 12^ However, for cutoff −10 HU, higher LmS attenuation had higher mortality risk even after adjustment for clinical and MPI variables, implying the potential residual confounding factors not recorded in our registry such as cirrhosis, hepatocellular carcinoma, or other organ dysfunction.^8, 24^ The ratio of liver attenuation to spleen (L2S) attenuation was not considered in this study due to the large variation of cutoffs reported in literature.^28–30^ Information on other liver health specific risk factors, for example, alcohol use, hepatocellular carcinoma, etc. was not available as our registry is focused on cardiovascular risk testing. Our study was retrospective in nature, and further prospective studies are needed to confirm our findings and evaluate the impact of interventions based on these findings. Finally, though the generalizability of our findings to populations undergoing MPI at different sites with different scanner characteristics or acquisition protocols warrants further investigation was validated, it is possible that site-, population- and scanner-specific refinement can offer even better risk stratification and patient management due to potential variation caused by patient characteristics and imaging protocols.

In conclusion, AI-driven HS detection from standard cardiac CTAC scans obtained during MPI provides an efficient and automated method for identifying patients at elevated cardiometabolic risk without additional imaging or physician involvement. The integrated liver risk index further improves prognostication, potentially guiding clinical decisions and targeted interventions to mitigate risk and improve patient outcomes.

## List of abbreviations

ACM: all-cause mortality
AI: artificial intelligence
BMI: body mass index
CAC: coronary artery calcium
CAD: coronary artery disease
CTAC: computed tomography attenuation correction
HU: Hounsfield unit
LVEF: left ventricular ejection fraction
MPI: myocardial perfusion imaging
PET: positron emission tomography
SPECT: single-photon emission computed tomography
TPD: total perfusion deficit

## Contributors

JY co-designed the study, developed the algorithms/models, conducted data processing/experiments/analysis, and co-wrote the manuscript. KP, RM and AM co-wrote the manuscript and contributed materials, clinical expertise, and technical expertise. PJS co-designed the study, provided the overall guidance and study funding, co-wrote the manuscript, and contributed materials, clinical expertise, and technical expertise. AS, WH, ND, ML, JZ, WZ, JXL, GR, VB, LS, MT, EA, IS, RP, MA, TDR, AJE, AF, EJM, WA, SK, VL, SM, VC, PC, SW, AK, LW, DB, DD, and MDC contributed materials, clinical expertise, and technical expertise. All authors critically revised the manuscript and contributed to its formation. JY, KP, RM, AM, AS, and PJS directly accessed and verified the data in the study. All authors had full access to all the data in the study, accept the final responsibility to submit for publication and take responsibility for the contents of the manuscript.

## Declaration of interests

KKP reports an institutional research grant from Jubilant DraxImage and research support from American College of Cardiology Geriatric Cardiology council. RJM has received grant funding and consulting fees from Pfizer as well as grant funding from Alberta Innovates. AMM has received consulting fees from APQ Health. TDR has received research grant support from GE Healthcare and Advanced Accelerator Applications. AJE has received a speaker’s fee from Ionetix, consulting fees from W. L. Gore & Associates and Artrya, authorship fees from Wolters Kluwer Healthcare—UpToDate, and served on scientific advisory boards for Axcellant and Canon Medical Systems USA; his institution has grants/grants pending from Alexion, Attralus, BridgeBio, Canon Medical Systems USA, GE HealthCare, Intellia Therapeutics, Ionis Pharmaceuticals, Neovasc, Pfizer, Roche Medical Systems, Synektik, and W. L. Gore & Associates. EJM has received grant support and serves as a consultant for GE Healthcare. DD, DB, and PS have equity interest in APQ Health. Inc. DB and PS participate in software royalties for QPS software at Cedars-Sinai Medical Center. DB also served as a consultant for GE Healthcare. PS has received research grant support from Siemens Medical Systems and consulting fees from Synektik S.A. The remaining authors have declared no competing interests.

## Data sharing

All implementations of the proposed segmentation and quantification approach primarily utilized Python-based libraries, including Python version 3.11.5, XGBoost version 2.1.1 (https://xgboost.readthedocs.io/en/latest/index.html), Scikit-image version 0.20.0 (https://scikit-image.org/), OpenCV version 4.6.0 (https://opencv.org/), and SciPy version 1.11.3 (https://scipy.org/). The TotalSegmentator v2 (https://github.com/wasserth/TotalSegmentator), and CAC segmentation model were implemented in PyTorch (https://pytorch.org/). All statistical analyses were performed using packages in RStudio 4.3.2, with the following R libraries: dplyr, tidyr, readxl, stringr, corrplot, survival, survminer, VennDiagram, gtsummary, gt, forestmodel, tidyverse, tidytidbits, adjustedCurves, reshape2, pylr, readxl, compare, survcomp and survivalAnalysis. All experiments were conducted on a desktop system running Windows 10 Pro 64-bit, equipped with 256GB RAM, an AMD Ryzen Threadripper 3950X 24-Core Processor, and an NVIDIA TITAN RTX 24GB GPU. The source code is available upon reasonable written request and can be accessed from the date of publication.

## Acknowledgements

This research was supported in part by grant R35HL161195 from the National Heart, Lung, and Blood Institute of the National Institutes of Health and R01EB034586 from the National Institute of Biomedical Imaging and Bioengineering of the National Institutes of Health. The content is solely the responsibility of the authors and does not necessarily represent the official views of the National Institutes of Health.

## Notes

### Author Declarations

The study followed the Declaration of Helsinki, with institutional review board (IRB) approval obtained from each participating institution. IRB of Cedars-Sinai Medical Center in Los Angeles, California, granted overall study approval.

### Summary of Updates

This version of the manuscript has been revised to correct an error in the Funding section. The funding information has now been updated accurately.

