## Supplemental Material for "AI-based Hepatic Steatosis Detection and Integrated Hepatic Assessment from Cardiac CT Attenuation Scans Enhances All-cause Mortality Risk Stratification: A Multi-center Study"

### Supplementary appendix

### Supplementary Methods

We developed and externally validated an eXtreme gradient boosting (XGBoost) decision tree model to combine hepatic measures and three demographic variables which were widely available and modified the association of hepatic steatosis with mortality to derive a liver risk index (LIRI) for integrated liver assessment, and the all-cause mortality status was used as ground truth label for training the model with a supervised binary classification task. Patient cases from one positron emission tomography (PET) site (Brigham and Women's Hospital (BWH)) and one single-photon emission computed tomography (SPECT) site (University of Calgary (CALGARY)) were used as external testing site data for evaluating prognostic value of LIRI while the cases from the other four PET sites (Cedars-Sinai Medical Center (CSMC), Intermountain Healthcare (ITMT), Mayo Clinic (MAYO), and University of Naples Federico II (NAPLES)) and the other three SPECT sites (Yale University (YALE), Columbia University (COLUMBIA), University of Ottawa (OTTAWA)) were used as model development dataset.

#### Data preprocessing

The 0.5th and 99.5th percentiles of numerical features were calculated over the model development dataset to truncate the features within these bounds to avoid potential outliers. The numerical features were then normalized to fall between 0 and 1. Subsequently, the mean and standard deviation of the numerical features over the model development dataset were computed to standardize the features, ensuring they have a zero mean and unit variance.

#### External testing scheme

To validate the generalizability of XGBoost models across sites, we trained and validated XGBoost models in an external testing scheme (Supplementary Figure 1).[1] Patients from four PET sites (CSMC, ITMT, MAYO and NAPLES) and three SPECT sites (YALE, COLUMBIA, and OTTAWA) were used as the development dataset to train the XGBoost model, while patients from the remaining PET site (BWH) and the remaining SPECT site (CALGARY) served as the external testing site dataset.

Hyperparameter tuning and selection

The development data was randomly and equally split into five non-overlapping folds. For each of the five folds, it was used as the validation fold, while the remaining four folds were used as the training data to train a model. During hyperparameter tuning (Supplementary Figure 1-A), each candidate hyperparameter configuration was used to train a model on the training data and then evaluated for area under receiver operating characteristic curve (AUC) on the corresponding validation data. The binary logistic loss function was used. The average AUC across all five validation folds was used to select the best-performing hyperparameter configuration. See Supplementary Table 2 for the searching space and the final configuration of the hyperparameter selection.

Model training

During model training (Supplementary Figure 1-B), the development data was randomly split into training (80%) and validation (20%) sets. The training data and the optimally selected hyperparameters were used to train an XGBoost model, while the validation data was used to select the number of trees for prediction on the external testing data. We repeated the model training twice with different random splits of the development data to obtain two models for prediction on the external testing data. In total, two XGBoost models were selected.

Inference and external testing

Both the external testing sites (i.e., BWH and CALGARY) were used for evaluating the prognostic value of LIRI produced by the model. During testing, the two models were used to make predictions for the external testing site data (Supplementary Figure 1-C), with the final prediction being the maximum of the two predictions.

### Supplementary Tables

**Supplementary Table 1. Overview of key imaging acquisition parameters.**

| Modality | Site | kVp | Tube current (mAs) | Breathing mode | Spacing (mm) | | | Dimensionality | | |
|---|---|---|---|---|---|---|---|---|---|---|
| | | | | | x | y | z | x | y | z |
| SPECT/CT | Yale | 120 | 20-150 | Normal breathing | 0.98 | 0.98 | 2.5-5 | 512 | 512 | 25-91 |
| | Calgary | 120 | 20-300 | End-expiratory breath hold | 0.49-0.98 | 0.49-0.98 | 2.5-5 | 512 | 512 | 45-157 |
| | Columbia | 90-120 | 31-125 | Normal breathing | 1.17 | 1.17 | 3 | 512 | 512 | 54-166 |
| | Ottawa | 130 | 24-53 | End-expiratory breath hold | 0.98 | 0.98 | 5 | 512 | 512 | 28-84 |
| PET/CT | CSMC | 100 | 11-13 | Normal breathing | 1.37 | 1.37 | 3 | 512 | 512 | 39-75 |
| | BWH | 80-140 | 10-300 | Normal breathing | 0.98-1.37 | 0.98-1.37 | 2.5-5 | 512 | 512 | 24-79 |
| | Mayo | 120-140 | 20-180 | Normal breathing | 1.37 | 1.37 | 3.75 | 512 | 512 | 27-47 |
| | ITMT | 120-130 | 16-407 | Normal breathing | 1.37 | 1.37 | 2 | 512 | 512 | 82-111 |
| | NAPLES | 120-140 | 35-260 | Normal breathing | 1.17 | 1.17 | 3 | 512 | 512 | 59-60 |

kVp – kilovoltage peak, SPECT – single-photon emission computed tomography, CT – computed tomography, PET – positron emission tomography, Yale – Yale University, Calgary – University of Calgary, Columbia – Columbia University, Ottawa – University of Ottawa, CSMC – Cedars-Sinai Medical Center, BWH – Brigham and Women's Hospital, Mayo – Mayo Clinic, ITMT – Intermountain Healthcare

**Supplementary Table 2. Searching space for hyperparameter tuning and final selected hyperparameters for training eXtreme Gradient Boosting (XGBoost) model.**

| Hyperparameter | Searching space | Selected value |
|---|---|---|
| "**max_depth**": maximum depth of a tree with larger value for more complex model | 3, 5, 7 | 5 |
| "**learning_rate**": step size or scaling coefficient with larger value for more contribution of a tree prediction | 0.01, 0.05, 0.3 | 0.3 |
| "**n_estimators**": number of gradient boost trees to use in an XGBoost model with larger value for more complex model | 100, 300, 500 | 100 |
| "**subsample**": sampling ratio of training instances for training each gradient boost tree with larger value for less biased estimation | 0.5, 0.7, 0.9 | 0.7 |
| "**colsample_bytree**": sampling ratio of features for training each gradient boost tree with larger value for more complex model | 0.5, 0.7, 0.9 | 0.9 |
| "**scale_pos_weight**": scaling coefficient of loss for positive instances with larger value for focusing more on improving prediction performance over positive instances | 6, 7, 8 | 8 |
| "**gamma**": minimum loss reduction required for growing a gradient boost tree depth with larger value for shorter tree and less complex model | 0, 0.2, 1 | 0 |
| "**lambda**": coefficient of $L_2$ regularization on gradient boost tree weights with larger value for smaller model weights and less complex model | 0, 0.1, 1 | 1 |
| "**alpha**": coefficient of $L_1$ regularization on gradient boost tree weights with larger value for sparser weights and less complex model | 0, 0.1, 1 | 1 |

**Supplementary Table 3. Baseline characteristics for all participants stratified by perfusion imaging modality (SPECT/CT vs PET/CT).**

| Characteristics | Overall<br>N = 27039 | PET<br>N = 16740 | SPECT<br>N = 10299 | p-value |
|---|---|---|---|---|
| **Demographics** | | | | |
| Age [year] | 67 (58, 75) | 68 (59, 76) | 65 (57, 73) | **<0.0001** |
| age ≥ 60 years | 19333 (72%) | 12395 (74%) | 6938 (67%) | **<0.0001** |
| Sex | | | | **<0.0001** |
| Female | 11689 (43%) | 7068 (42%) | 4621 (45%) | |
| Male | 15350 (57%) | 9672 (58%) | 5678 (55%) | |
| BMI [kg/m$^2$] | 29.5 (25.5, 34.7) | 29.3 (25.4, 34.9) | 29.9 (25.8, 34.6) | **0.0152** |
| Race | | | | **<0.0001** |
| American Indian | 92 (0.3%) | 73 (0.4%) | 19 (0.2%) | |
| Asian | 580 (2.1%) | 414 (2.5%) | 166 (1.6%) | |
| Black | 2648 (9.8%) | 1452 (8.7%) | 1196 (12%) | |
| Pacific Islander | 149 (0.6%) | 129 (0.8%) | 20 (0.2%) | |
| White | 17186 (64%) | 13681 (82%) | 3505 (34%) | |
| Unknown | 6384 (24%) | 991 (5.9%) | 5393 (52%) | |
| **Medical Conditions** | | | | |
| Hypertension | 20096 (74%) | 13732 (82%) | 6364 (62%) | **<0.0001** |
| Diabetes mellitus | 8950 (33%) | 6072 (36%) | 2878 (28%) | **<0.0001** |
| Dyslipidemia | 17645 (65%) | 12434 (74%) | 5211 (51%) | **<0.0001** |
| Family history of coronary artery disease | 8030 (30%) | 5502 (33%) | 2528 (25%) | **<0.0001** |
| Smoking | 4644 (17%) | 3155 (19%) | 1489 (14%) | **<0.0001** |
| BMI ≥ 30 kg/m$^2$ | 12889 (48%) | 7775 (46%) | 5114 (50%) | **<0.0001** |
| Stress TPD ≥ 5% | 12341 (46%) | 8325 (50%) | 4016 (39%) | **<0.0001** |
| LVEF < 50% | 4864 (18%) | 3212 (19%) | 1652 (16%) | **<0.0001** |
| MFR < 2* | 6055 (36%) | 6055 (36%) | 0 (NA%) | **<0.0001** |
| **Imaging Measures** | | | | |
| CAC score [AU]# | 2.08 (0.00, 2.95) | 2.21 (0.00, 3.01) | 1.80 (0.00, 2.84) | **<0.0001** |
| Stress TPD [%] | 4 (2, 10) | 5 (2, 12) | 3 (1, 8) | **<0.0001** |
| LVEF [%] | 66 (55, 75) | 68 (55, 76) | 64 (55, 72) | **<0.0001** |
| MFR* | 2.26 (1.77, 2.82) | 2.26 (1.77, 2.82) | NA (NA, NA) | |
| **Hepatic Measures** | | | | |
| Liver attenuation [HU] | 48 (40, 54) | 47 (40, 53) | 50 (42, 56) | **<0.0001** |
| Liver SD [HU] | 44 (31, 56) | 40 (25, 56) | 47 (39, 56) | **<0.0001** |
| Liver entropy [bit] | 7.49 (6.97, 7.85) | 7.35 (6.67, 7.83) | 7.60 (7.32, 7.86) | **<0.0001** |
| Liver CoV | 0.89 (0.61, 1.30) | 0.85 (0.53, 1.33) | 0.94 (0.73, 1.27) | **<0.0001** |
| LmS attenuation [HU] | 13 (7, 18) | 14 (8, 19) | 11 (5, 17) | **<0.0001** |
| LmS SD [HU] | -2.3 (-5.3, 0.0) | -2.2 (-4.3, -0.2) | -2.9 (-8.1, 0.5) | **<0.0001** |
| LmS entropy [bit] | -0.08 (-0.17, 0.00) | -0.08 (-0.15, -0.01) | -0.08 (-0.22, 0.02) | **<0.0001** |
| LmS CoV | -0.30 (-0.62, -0.14) | -0.32 (-0.67, -0.15) | -0.27 (-0.54, -0.12) | **<0.0001** |
| Hepatic steatosis | 6579 (24%) | 4357 (26%) | 2222 (22%) | **<0.0001** |
| **Outcome** | | | | |
| Death | 4836 (18%) | 4200 (25%) | 636 (6.2%) | **<0.0001** |
| Death follow up [year] | 3.58 (1.86, 5.15) | 4.57 (2.59, 5.73) | 2.32 (1.42, 3.65) | **<0.0001** |

Values are presented as N (%) or median (IQ1, IQ3). Bold indicates statistical significance with p<0.05. Abnormal stress TPD was defined as stress TPD ≥ 5% and abnormal LVEF was defined as LVEF ≥ 50%. * only PET/CT patients had MFR measurements. # logarithmic transformed CAC score via log10(CAC score+1).

SPECT/CT – single-photon emission computed tomography/computed tomography, PET/CT – positron emission tomography/computed tomography, BMI – body mass index, CAD – coronary artery disease, TPD – total perfusion deficit, LVEF – left ventricular ejection fraction, CAC – coronary artery calcium, AU – Agatston unit, HU – Hounsfield unit, CoV – coefficient of variation, LmS attenuation – difference of liver attenuation minus spleen attenuation, LmS CoV – difference of liver CoV minus spleen CoV, LmS SD – difference of liver SD minus spleen SD, LmS entropy – difference of liver entropy minus spleen entropy, MFR – myocardial flow reserve

**Supplementary Table 4. Hepatic measures for all participants stratified by different characteristics.**

| Stratified by sex | | | | |
|---|---|---|---|---|
| Measures | Overall<br>N = 27039 | Female<br>N = 11689 | Male<br>N = 15350 | p-value |
| Liver attenuation [HU] | 48 (40, 54) | 50 (42, 56) | 47 (39, 53) | **<0.0001** |
| Liver CoV | 0.89 (0.61, 1.30) | 0.78 (0.55, 1.11) | 0.99 (0.68, 1.43) | **<0.0001** |
| Liver entropy [bit] | 7.49 (6.97, 7.85) | 7.34 (6.86, 7.70) | 7.60 (7.07, 7.94) | **<0.0001** |
| Liver SD [HU] | 44 (31, 56) | 40 (29, 51) | 47 (33, 60) | **<0.0001** |
| LmS attenuation [HU] | 13 (7, 18) | 13 (8, 19) | 12 (6, 18) | **<0.0001** |
| LmS CoV | -0.30 (-0.62, -0.14) | -0.29 (-0.54, -0.15) | -0.31 (-0.69, -0.13) | **<0.0001** |
| LmS entropy [bit] | -0.08 (-0.17, 0.00) | -0.09 (-0.18, -0.01) | -0.07 (-0.16, 0.01) | **<0.0001** |
| LmS SD [HU] | -2.3 (-5.3, 0.0) | -2.5 (-5.2, -0.4) | -2.2 (-5.6, 0.3) | **<0.0001** |
| Hepatic steatosis | 6579 (24%) | 2402 (21%) | 4177 (27%) | **<0.0001** |
| Death | 4836 (18%) | 1900 (16%) | 2936 (19%) | **<0.0001** |
| Death follow up [year] | 3.58 (1.86, 5.15) | 3.65 (1.98, 5.18) | 3.53 (1.79, 5.11) | **<0.0001** |
| Stratified by BMI | | | | |
| Measures | Overall<br>N = 27039 | BMI < 30 $kg/m^2$<br>N = 14150 | BMI ≥ 30 $kg/m^2$<br>N = 12889 | p-value |
| Liver attenuation [HU] | 48 (40, 54) | 52 (46, 57) | 44 (34, 50) | **<0.0001** |
| Liver CoV | 0.89 (0.61, 1.30) | 0.78 (0.55, 1.07) | 1.07 (0.73, 1.59) | **<0.0001** |
| Liver entropy [bit] | 7.49 (6.97, 7.85) | 7.37 (6.90, 7.74) | 7.62 (7.09, 7.93) | **<0.0001** |
| Liver SD [HU] | 44 (31, 56) | 41 (29, 52) | 48 (33, 59) | **<0.0001** |
| LmS attenuation [HU] | 13 (7, 18) | 13 (8, 18) | 12 (4, 18) | **<0.0001** |
| LmS CoV | -0.30 (-0.62, -0.14) | -0.30 (-0.56, -0.16) | -0.31 (-0.69, -0.10) | **0.0381** |
| LmS entropy [bit] | -0.08 (-0.17, 0.00) | -0.10 (-0.18, -0.02) | -0.05 (-0.15, 0.03) | **<0.0001** |
| LmS SD [HU] | -2.3 (-5.3, 0.0) | -2.8 (-5.7, -0.6) | -1.8 (-4.8, 0.7) | **<0.0001** |
| Hepatic steatosis | 6579 (24%) | 1586 (11%) | 4993 (39%) | **<0.0001** |
| Death | 4836 (18%) | 2992 (21%) | 1844 (14%) | **<0.0001** |
| Death follow up [year] | 3.58 (1.86, 5.15) | 3.54 (1.77, 5.13) | 3.63 (1.97, 5.17) | **<0.0001** |
| Stratified by age | | | | |
| Measures | Overall<br>N = 27039 | < 60 years old<br>N = 7706 | ≥ 60 years old<br>N = 19333 | p-value |
| Liver attenuation [HU] | 48 (40, 54) | 48 (37, 55) | 49 (41, 54) | **<0.0001** |
| Liver CoV | 0.89 (0.61, 1.30) | 0.91 (0.60, 1.36) | 0.89 (0.61, 1.28) | **0.0012** |
| Liver entropy [bit] | 7.49 (6.97, 7.85) | 7.50 (6.96, 7.84) | 7.49 (6.97, 7.85) | 0.3798 |
| Liver SD [HU] | 44 (31, 56) | 44 (30, 56) | 44 (31, 56) | 0.2198 |
| LmS attenuation [HU] | 13 (7, 18) | 11 (4, 17) | 13 (8, 19) | **<0.0001** |
| LmS CoV | -0.30 (-0.62, -0.14) | -0.26 (-0.53, -0.10) | -0.32 (-0.65, -0.16) | **<0.0001** |
| LmS entropy [bit] | -0.08 (-0.17, 0.00) | -0.08 (-0.17, 0.01) | -0.08 (-0.17, 0.00) | 0.0977 |
| LmS SD [HU] | -2.3 (-5.3, 0.0) | -2.4 (-5.4, 0.0) | -2.3 (-5.3, 0.0) | 0.1847 |
| Hepatic steatosis | 6579 (24%) | 2342 (30%) | 4237 (22%) | **<0.0001** |

| | | | | |
|---|---|---|---|---|
| Death | 4836 (18%) | 723 (9.4%) | 4113 (21%) | **<0.0001** |
| Death follow up [year] | 3.58 (1.86, 5.15) | 3.67 (1.82, 5.25) | 3.56 (1.88, 5.11) | **0.0341** |
| Stratified by race | | | | |
| Measures | Overall<br>N = 19834 | Black<br>N = 2648 | White<br>N = 17186 | p-value |
| Liver attenuation [HU] | 48 (40, 54) | 50 (42, 56) | 48 (40, 53) | **<0.0001** |
| Liver CoV | 0.92 (0.59, 1.36) | 0.94 (0.63, 1.40) | 0.92 (0.59, 1.36) | **0.002** |
| Liver entropy [bit] | 7.51 (6.84, 7.92) | 7.61 (7.04, 8.01) | 7.48 (6.80, 7.90) | **<0.0001** |
| Liver SD [HU] | 44 (28, 59) | 48 (32, 63) | 44 (28, 58) | **<0.0001** |
| LmS attenuation [HU] | 13 (7, 19) | 15 (9, 21) | 13 (7, 18) | **<0.0001** |
| LmS CoV | -0.33 (-0.70, -0.15) | -0.38 (-0.93, -0.17) | -0.33 (-0.67, -0.15) | **<0.0001** |
| LmS entropy [bit] | -0.08 (-0.17, 0.00) | -0.10 (-0.21, 0.00) | -0.08 (-0.17, 0.00) | **<0.0001** |
| LmS SD [HU] | -2.4 (-5.5, -0.2) | -3.3 (-8.4, -0.2) | -2.3 (-5.2, -0.2) | **<0.0001** |
| Hepatic steatosis | 4999 (25%) | 538 (20%) | 4461 (26%) | **<0.0001** |
| Death | 4084 (21%) | 560 (21%) | 3524 (21%) | 0.4463 |
| Death follow up [year] | 4.02 (2.00, 5.32) | 3.55 (1.73, 6.19) | 4.12 (2.03, 5.27) | 0.1671 |
| Stratified by diabetes | | | | |
| Measures | Overall<br>N = 27039 | Non-diabetic<br>N = 18089 | Diabetic<br>N = 8950 | p-value |
| Liver attenuation [HU] | 48 (40, 54) | 50 (42, 55) | 45 (37, 52) | **<0.0001** |
| Liver CoV | 0.89 (0.61, 1.30) | 0.87 (0.60, 1.24) | 0.96 (0.63, 1.45) | **<0.0001** |
| Liver entropy [bit] | 7.49 (6.97, 7.85) | 7.48 (7.00, 7.82) | 7.52 (6.86, 7.89) | **0.0487** |
| Liver SD [HU] | 44 (31, 56) | 44 (31, 55) | 45 (28, 58) | 0.066 |
| LmS attenuation [HU] | 13 (7, 18) | 13 (7, 18) | 12 (5, 18) | **<0.0001** |
| LmS CoV | -0.30 (-0.62, -0.14) | -0.30 (-0.60, -0.15) | -0.30 (-0.66, -0.11) | **0.0157** |
| LmS entropy [bit] | -0.08 (-0.17, 0.00) | -0.08 (-0.17, 0.00) | -0.07 (-0.16, 0.01) | **<0.0001** |
| LmS SD [HU] | -2.3 (-5.3, 0.0) | -2.4 (-5.5, -0.1) | -2.1 (-4.9, 0.3) | **<0.0001** |
| Hepatic steatosis | 6579 (24%) | 3658 (20%) | 2921 (33%) | **<0.0001** |
| Death | 4836 (18%) | 2771 (15%) | 2065 (23%) | **<0.0001** |
| Death follow up [year] | 3.58 (1.86, 5.15) | 3.60 (1.88, 5.11) | 3.56 (1.84, 5.21) | 0.9971 |
| Stratified by perfusion | | | | |
| Measures | Overall<br>N = 27039 | Stress TPD < 5%<br>N = 14698 | Stress TPD ≥ 5%<br>N = 12341 | p-value |
| Liver attenuation [HU] | 48 (40, 54) | 49 (41, 55) | 47 (40, 53) | **<0.0001** |
| Liver CoV | 0.89 (0.61, 1.30) | 0.93 (0.67, 1.33) | 0.85 (0.55, 1.26) | **<0.0001** |
| Liver entropy [bit] | 7.49 (6.97, 7.85) | 7.56 (7.14, 7.89) | 7.38 (6.72, 7.78) | **<0.0001** |
| Liver SD [HU] | 44 (31, 56) | 46 (35, 58) | 41 (26, 54) | **<0.0001** |
| LmS attenuation [HU] | 13 (7, 18) | 13 (7, 18) | 12 (6, 18) | **0.004** |
| LmS CoV | -0.30 (-0.62, -0.14) | -0.33 (-0.67, -0.16) | -0.27 (-0.55, -0.12) | **<0.0001** |
| LmS entropy [bit] | -0.08 (-0.17, 0.00) | -0.09 (-0.18, -0.01) | -0.06 (-0.15, 0.02) | **<0.0001** |
| LmS SD [HU] | -2.3 (-5.3, 0.0) | -2.9 (-6.3, -0.4) | -1.7 (-4.3, 0.4) | **<0.0001** |
| Hepatic steatosis | 6579 (24%) | 3393 (23%) | 3186 (26%) | **<0.0001** |
| Death | 4836 (18%) | 1828 (12%) | 3008 (24%) | **<0.0001** |

| | | | | |
|---|---|---|---|---|
| Death follow up [year] | 3.58 (1.86, 5.15) | 3.67 (1.94, 5.11) | 3.46 (1.78, 5.20) | **0.0011** |
| Stratified by myocardial flow reserve | | | | |
| Measures | Overall<br>N = 16740 | Non-CMD (MFR ≥ 2)<br>N = 10685 | CMD (MFR < 2)<br>N = 6055 | p-value |
| Liver attenuation [HU] | 47 (40, 53) | 48 (40, 53) | 47 (39, 53) | **0.0001** |
| Liver CoV | 0.85 (0.53, 1.33) | 0.89 (0.55, 1.36) | 0.78 (0.50, 1.26) | **<0.0001** |
| Liver entropy [bit] | 7.35 (6.67, 7.83) | 7.43 (6.75, 7.86) | 7.19 (6.59, 7.79) | **<0.0001** |
| Liver SD [HU] | 40 (25, 56) | 42 (27, 56) | 36 (24, 54) | **<0.0001** |
| LmS attenuation [HU] | 14 (8, 19) | 14 (8, 19) | 14 (8, 19) | **0.0096** |
| LmS CoV | -0.32 (-0.67, -0.15) | -0.33 (-0.66, -0.15) | -0.32 (-0.69, -0.14) | 0.4169 |
| LmS entropy [bit] | -0.08 (-0.15, -0.01) | -0.08 (-0.14, -0.01) | -0.08 (-0.16, 0.00) | 0.4042 |
| LmS SD [HU] | -2.2 (-4.3, -0.2) | -2.3 (-4.4, -0.4) | -2.0 (-4.2, 0.0) | **<0.0001** |
| Hepatic steatosis | 4357 (26%) | 2751 (26%) | 1606 (27%) | 0.2709 |
| Death | 4200 (25%) | 1776 (17%) | 2424 (40%) | **<0.0001** |
| Death follow up [year] | 4.6 (2.6, 5.7) | 4.7 (3.1, 5.8) | 4.1 (1.9, 5.7) | **<0.0001** |

Values are presented as N (%) or median (IQ1, IQ3)
BMI – body mass index, CAD – coronary artery disease, TPD – total perfusion deficit, LVEF – left ventricular ejection fraction, CAC – coronary artery calcium, CoV – coefficient of variation, LmS attenuation – difference of liver attenuation minus spleen attenuation, LmS CoV – difference of liver CoV minus spleen CoV, LmS standard deviation – difference of liver standard deviation minus spleen standard deviation, LmS entropy – difference of liver entropy minus spleen entropy, MFR – myocardial flow reserve, CMD – coronary microvascular disease, HU – Hounsfield unit

**Supplementary Table 5. Maximally selected rank statistics-based optimal cutoffs for our proposed liver and spleen measures in entire population for all-cause mortality prediction in MPI cohort of 27039 patients.** For the liver attenuation and the difference of liver attenuation minus spleen attenuation (LmS attenuation), we used the cutoff 40 HU and -10 HU from existing literatures, respectively.[2, 3] We used the maximally selected rank statistic-based cutoff for other hepatic measures.[4]

| | Cutoff | Concordance-index [95% CI] |
|---|---|---|
| Liver attenuation [HU] | 40 | 0.50 [0.49, 0.51] |
| Liver CoV | 0.5752 | 0.53 [0.52, 0.53] |
| LmS attenuation [HU] | -10 | 0.52 [0.48, 0.53] |
| LmS CoV | 0.0136 | 0.50 [0.49, 0.51] |
| LmS entropy [bit] | 0.0449 | 0.52 [0.51, 0.53] |
| LmS SD [HU] | 0.2678 | 0.48 [0.47, 0.53] |

CI – confidence interval, HU – Hounsfield unit, CoV – coefficient of variation, LmS – liver metric minus spleen metric, SD – standard deviation

**Supplementary Table 6. Comparison between adjusted hazard ratios of hepatic measure in males and females**

| In different sex populations | | |
|---|---|---|
| Factor | Hazard ratio (HR) [95% CI], p value* | |
| | Female<br>N=11689, 1900 events | Male<br>N=15350, 2936 events |
| HSIC-based hepatic steatosis | 1.02 [0.89, 1.18], 0.7459 | **1.21 [1.1, 1.34], 0.0001** |
| High liver CoV | **0.87 [0.78, 0.97], 0.0093** | **0.8 [0.73, 0.88], <0.0001** |
| High LmS CoV | 1.08 [0.9, 1.31], 0.3996 | 1.05 [0.91, 1.2], 0.5199 |
| High LmS entropy | 1.14 [0.94, 1.38], 0.1895 | **1.21 [1.05, 1.39], 0.0096** |
| High LmS SD | **1.28 [1.08, 1.5], 0.0038** | **1.19 [1.05, 1.36], 0.0059** |
| In different age populations | | |
| Factor | Hazard ratio (HR) [95% CI], p value* | |
| | age < 60<br>N=7706, 723 events | age ≥ 60<br>N=19333, 4113 events |
| HSIC-based hepatic steatosis | 0.98 [0.79, 1.2], 0.8216 | **1.18 [1.08, 1.28], 0.0003** |
| High liver CoV | 1.01 [0.85, 1.21], 0.9009 | **0.81 [0.75, 0.87], <0.0001** |
| High LmS CoV | 1.04 [0.81, 1.33], 0.753 | 1.08 [0.96, 1.22], 0.2013 |
| High LmS entropy | 1.07 [0.81, 1.42], 0.6141 | **1.2 [1.06, 1.36], 0.0051** |
| High LmS SD | **1.72 [1.34, 2.2], <0.0001** | **1.15 [1.03, 1.28], 0.0126** |
| In different BMI populations | | |
| Factor | Hazard ratio (HR) [95% CI], p value* | |
| | BMI ≥ 30 $kg/m^2$<br>N=14150, 2992 events | BMI < 30 $kg/m^2$<br>N=12889, 1844 events |
| HSIC-based hepatic steatosis | 1.01 [0.9, 1.13], 0.8369 | **1.21 [1.07, 1.36], 0.0015** |
| High liver CoV | 0.91 [0.8, 1.02], 0.1051 | **0.86 [0.79, 0.93], 0.0004** |
| High LmS CoV | 1.04 [0.9, 1.2], 0.5909 | 1.1 [0.93, 1.3], 0.2603 |
| High LmS entropy | 1.03 [0.87, 1.23], 0.7337 | **1.3 [1.11, 1.51], 0.0009** |
| High LmS SD | **1.23 [1.05, 1.44], 0.0115** | **1.19 [1.05, 1.36], 0.007** |
| In different race populations | | |
| Factor | Hazard ratio (HR) [95% CI], p value* | |
| | Black<br>N=2648, 560 events | White<br>N=17186, 3524 events |
| HSIC-based hepatic steatosis | 0.91 [0.7, 1.17], 0.4497 | **1.21 [1.11, 1.33], <0.0001** |
| High liver CoV | 0.88 [0.72, 1.08], 0.2172 | **0.81 [0.75, 0.88], <0.0001** |
| High LmS CoV | 1.22 [0.88, 1.7], 0.2346 | 1.1 [0.97, 1.25], 0.1288 |
| High LmS entropy | 1.04 [0.72, 1.49], 0.8391 | **1.24 [1.08, 1.41], 0.0018** |
| High LmS SD | 1.21 [0.88, 1.66], 0.2502 | **1.21 [1.08, 1.36], 0.0012** |
| In patients with different diabetes status | | |
| Factor | Hazard ratio (HR) [95% CI], p value* | |
| | With diabetes<br>N=18089, 2771 events | No diabetes<br>N=8950, 2065 events |
| HSIC-based hepatic steatosis | 0.97 [0.86, 1.09], 0.6011 | **1.31 [1.17, 1.46], <0.0001** |
| High liver CoV | **0.87 [0.78, 0.96], 0.0079** | **0.82 [0.75, 0.9], <0.0001** |
| High LmS CoV | 1.06 [0.91, 1.24], 0.4479 | 1.06 [0.91, 1.24], 0.429 |

| | | |
|---|---|---|
| High LmS entropy | **1.25 [1.05, 1.48], 0.0123** | 1.14 [0.98, 1.33], 0.0959 |
| High LmS SD | **1.24 [1.06, 1.45], 0.0059** | **1.19 [1.04, 1.35], 0.0119** |
| In different perfusion populations | | |
| Factor | Hazard ratio (HR) [95% CI], p value* | |
| | Normal perfusion (stress TPD < 5%) N=14698, 1828 events | Abnormal perfusion (stress TPD ≥ 5%) N=12341, 3008 events |
| HSIC-based hepatic steatosis | 1.11 [0.97, 1.27], 0.1186 | **1.15 [1.04, 1.28], 0.0062** |
| High liver CoV | **0.8 [0.71, 0.9], 0.0002** | **0.87 [0.8, 0.94], 0.0009** |
| High LmS CoV | 0.91 [0.75, 1.11], 0.3524 | **1.14 [1, 1.31], 0.0455** |
| High LmS entropy | **1.25 [1.04, 1.51], 0.0191** | 1.15 [1, 1.33], 0.0576 |
| High LmS SD | **1.29 [1.1, 1.51], 0.002** | **1.18 [1.03, 1.34], 0.0138** |
| In different myocardial flow reserve populations (in PET population only) | | |
| Factor | Adjusted HR [95%], p-value* | |
| | MFR < 2 N=10685, 1776 events | MFR ≥ 2 N=6055, 2424 events |
| HSIC-based hepatic steatosis | 1.04 [0.93, 1.17], 0.448 | 1.14 [1, 1.3], 0.0535 |
| High liver CoV | **0.87 [0.79, 0.95], 0.0028** | **0.82 [0.73, 0.91], 0.0004** |
| High LmS CoV | **1.18 [1.01, 1.37], 0.0383** | 1.08 [0.9, 1.28], 0.4178 |
| High LmS entropy | **1.25 [1.06, 1.46], 0.0064** | 1.14 [0.94, 1.38], 0.1804 |
| High LmS SD | **1.24 [1.07, 1.42], 0.003** | **1.31 [1.11, 1.54], 0.0015** |

Bold indicates statistical significance.

Clinical and perfusion factors: modality, age, body mass index, hypertension, diabetes mellitus, dyslipidemia, family history of coronary artery disease, smoking, stress total perfusion deficit, left ventricle ejection fraction, and deep-learning-coronary artery calcium score. *Adjusted for clinical, perfusion factors and all the other hepatic quantifications. HR of age/BMI/log10(CAC score+1) was per unit increase. For modality, SPECT was the reference modality. LVEF ≥ 50%/Stress TPD<5%/low liver CoV/low LmS CoV/low LmS entropy/low LmS SD was reference group. For diabetes mellitus, dyslipidemia, family history, hypertension, and smoking, patients without them were the reference groups.

CI – confidence interval, BMI – body mass index, CAD – coronary artery disease, TPD – total perfusion deficit, LVEF – left ventricular ejection fraction, CAC – coronary artery calcium, CoV – coefficient of variation, LmS CoV – difference of liver CoV minus spleen CoV, LmS SD – difference of liver SD minus spleen SD, LmS entropy – difference of liver entropy minus spleen entropy, SPECT – single-photon emission computed tomography, PET – positron emission tomography, HSIC – hepatic steatosis imaging criterion

**Supplementary Table 7. Characteristics in external testing data of 7200 patients stratified by liver risk index (LIRI) categorized by a cutoff 0.63 which is determined by a maximally selected rank test.[4]**

| Characteristics | Overall<br>N = 7200 | High-risk (> 0.63)<br>N = 1537 | Low-risk (≤ 0.63)<br>N = 5663 | p-value |
|---|---|---|---|---|
| **Hepatic measures** | | | | |
| Liver attenuation | 52 (42, 58) | 50 (43, 57) | 52 (42, 59) | **0.0233** |
| Liver standard deviation | 30 (25, 44) | 29 (25, 40) | 31 (25, 45) | **<0.0001** |
| Liver entropy | 6.96 (6.62, 7.50) | 6.86 (6.63, 7.36) | 6.99 (6.62, 7.52) | **<0.0001** |
| Liver coefficient of variation | 0.61 (0.47, 0.87) | 0.59 (0.48, 0.82) | 0.61 (0.47, 0.89) | **0.0097** |
| LmS attenuation | 12 (6, 18) | 14 (8, 21) | 12 (6, 17) | **<0.0001** |
| LmS standard deviation | -2.3 (-4.4, -0.3) | -1.7 (-3.5, 0.3) | -2.5 (-4.6, -0.5) | **<0.0001** |
| LmS entropy | -0.10 (-0.18, -0.01) | -0.07 (-0.16, 0.01) | -0.11 (-0.19, -0.02) | **<0.0001** |
| LmS coefficient of variation | -0.22 (-0.38, -0.11) | -0.22 (-0.44, -0.11) | -0.22 (-0.37, -0.11) | 0.1309 |
| **Demographics** | | | | |
| Age | 66 (58, 74) | 80 (75, 84) | 63 (56, 70) | **<0.0001** |
| Sex | | | | **0.0001** |
| Female | 3295 (46%) | 636 (41%) | 2659 (47%) | |
| Male | 3905 (54%) | 901 (59%) | 3004 (53%) | |
| Body mass index | 29.0 (25.3, 33.8) | 26.8 (23.0, 31.0) | 29.7 (25.9, 34.5) | **<0.0001** |
| **Outcomes** | | | | |
| Death | 2059 (29%) | 775 (50%) | 1284 (23%) | **<0.0001** |
| Death follow up | 3.9 (2.0, 6.4) | 3.6 (1.7, 5.5) | 4.0 (2.1, 6.8) | **<0.0001** |

Median (IQR) or frequency (%). Bold indicates statistical significance. * CAC score is logarithmically transformed by log10(CAC score+1). LmS – liver minus spleen. LmS – difference of liver metric minus spleen metric

**Supplementary Table 8. Confusion matrices between hepatic steatosis (HS) and clinical/perfusion imaging variables.**

| | Age | |
|---|---|---|
| | < 60 years old | ≥ 60 years old |
| HS | 2342 (0.36) | 4237 (0.64) |
| No HS | 5364 (0.26) | **15096 (0.74)** |

| | Dyslipidemia | |
|---|---|---|
| | Yes | No |
| HS | **4348 (0.66)** | 2231 (0.34) |
| No HS | 13297 (0.65) | 7163 (0.35) |

| | Sex | |
|---|---|---|
| | Male | Female |
| HS | **4177 (0.63)** | 2402 (0.37) |
| No HS | 11173 (0.55) | 9287 (0.45) |

| | Family history of CAD | |
|---|---|---|
| | Yes | No |
| HS | 1889 (0.29) | 4690 (0.71) |
| No HS | 6141 (0.30) | **14319 (0.70)** |

| | Obesity | |
|---|---|---|
| | Yes | No |
| HS | **4993 (0.76)** | 1586 (0.24) |
| No HS | 7896 (0.39) | **12564 (0.61)** |

| | Smoking | |
|---|---|---|
| | Yes | No |
| HS | 1102 (0.17) | 5477 (0.83) |
| No HS | 3542 (0.17) | **16918 (0.83)** |

| | Hypertension | |
|---|---|---|
| | Yes | No |
| HS | **5178 (0.79)** | 1401 (0.21) |
| No HS | 14918 (0.73) | 5542 (0.27) |

| | Stress TPD | |
|---|---|---|
| | ≥ 5% | < 5% |
| HS | 3186 (0.48) | 3393 (0.52) |
| No HS | 9155 (0.45) | 11305 (0.55) |

| | Diabetes | |
|---|---|---|
| | Yes | No |
| HS | 2921 (0.44) | 3658 (0.56) |
| No HS | 6029 (0.29) | **14431 (0.71)** |

| | LVEF | |
|---|---|---|
| | < 50% | ≥ 50% |
| HS | 1075 (0.16) | 5504 (0.84) |
| No HS | 3789 (0.19) | **16671 (0.81)** |

| | CAC score | |
|---|---|---|
| | Non-zero | Zero |
| HS | **4281 (0.65)** | 2298 (0.35) |
| No HS | 14408 (0.70) | 6052 (0.30) |

| | MFR (in PET only) | |
|---|---|---|
| | < 2 | ≥ 2 |
| HS | 1606 (0.37) | 2751 (0.63) |
| No HS | 4449 (0.36) | **7934 (0.64)** |

Frequency (%). Bold indicates sensitivity or specificity no less than 0.60.
Age of 60 years was used to define young (less than 60) and old (no less than 60), body mass index (BMI) of 30 $kg/m^2$ was used to define obesity (no less than 30 $kg/m^2$) and non-obesity (less than 30 $kg/m^2$), stress total perfusion deficit (TPD) of 5% was used to define abnormal (no less than 5%) and normal (less than 5%), and left ventricle ejection fraction (LVEF) of 50% was used to define abnormal (less than 50%) and normal (no less than 50%). HS – hepatic steatosis, CAC – coronary artery calcium, MFR – myocardial flow reserve, PET – positron emission tomography

### Supplementary Figures

(A)

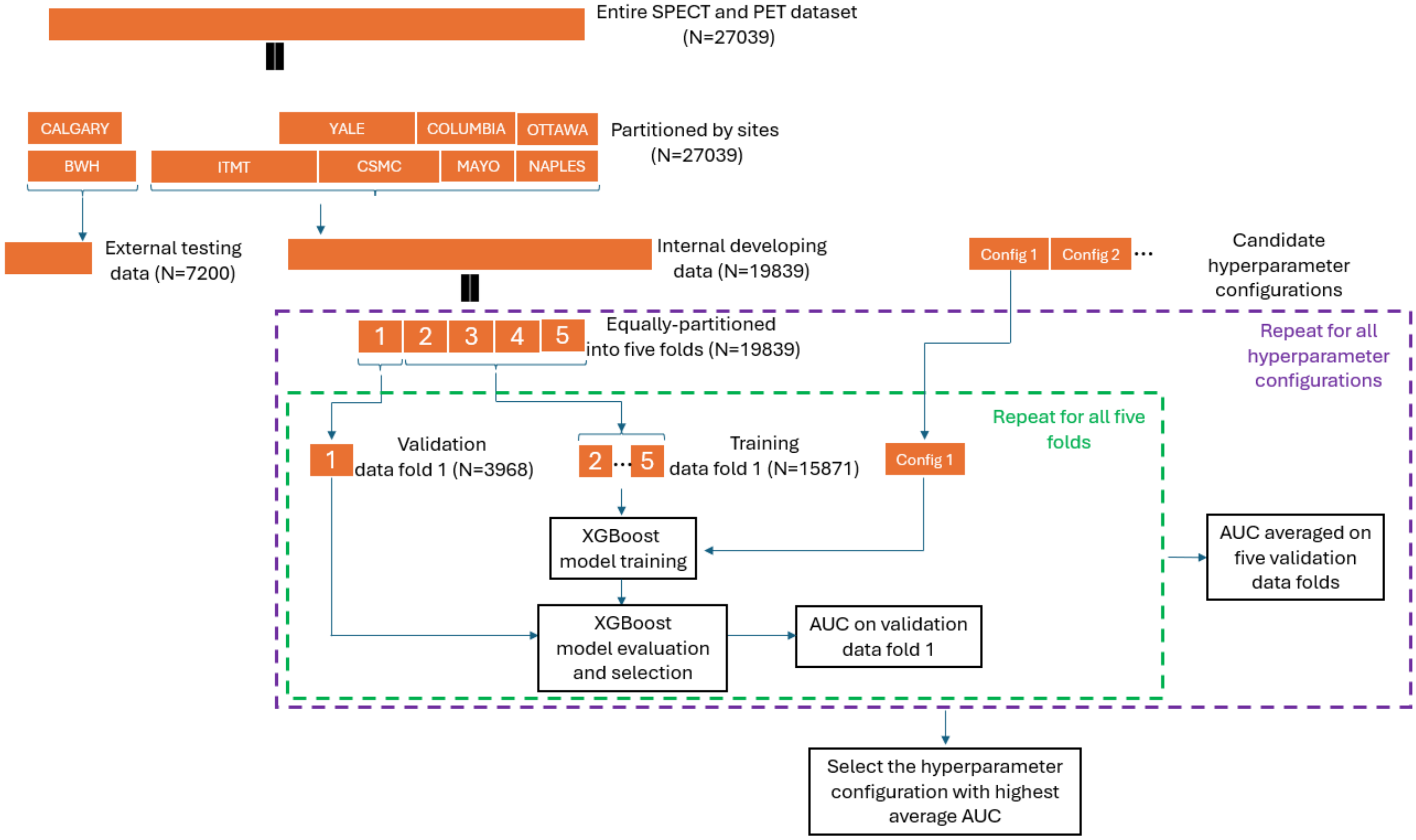

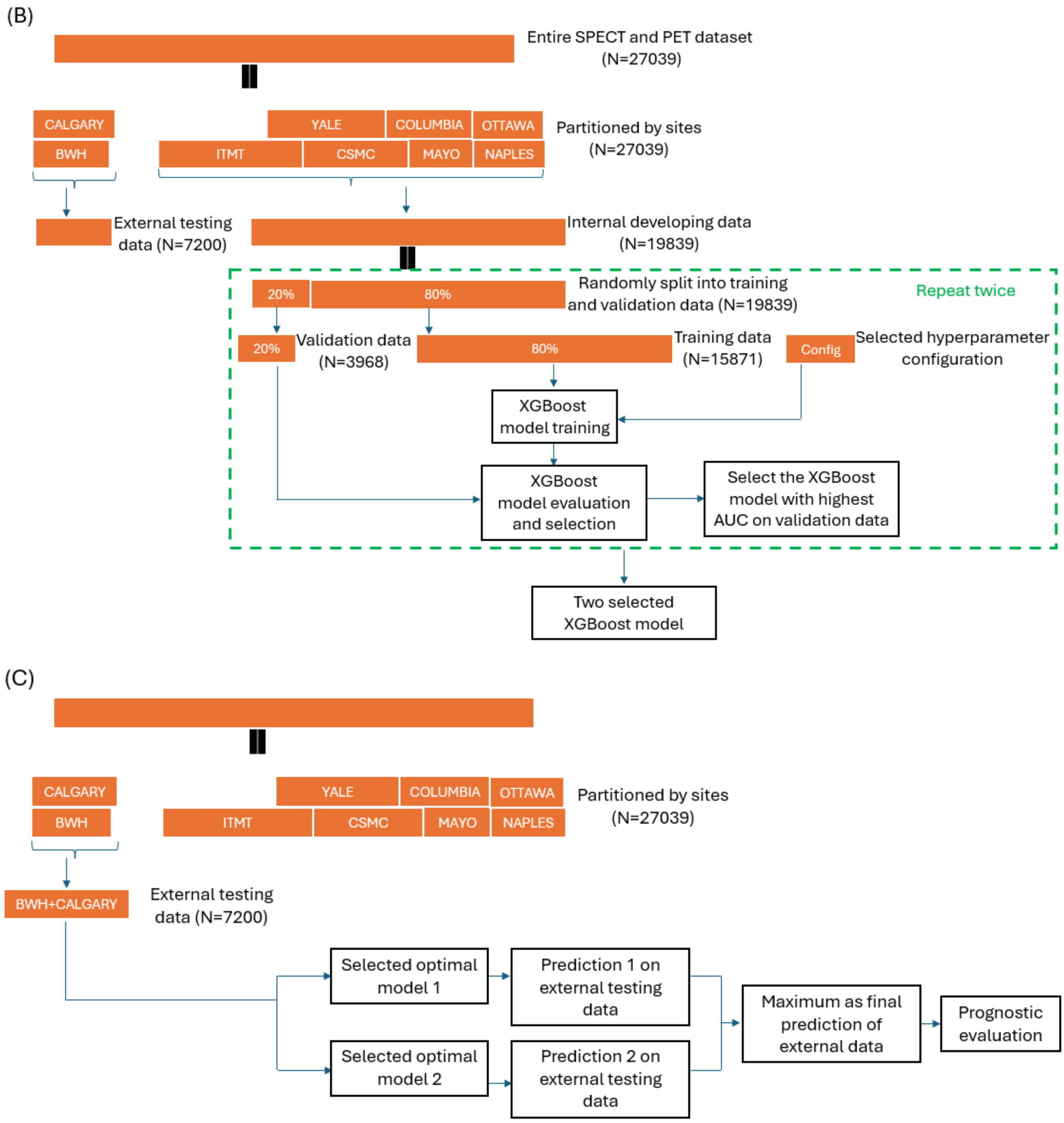


**Supplementary Figure 1. eXtreme Gradient Boosting (XGBoost) model development and validation in external testing scheme where patient cases from PET site Brigham and Women's Hospital (BWH) and SPECT site University of Calgary were used as external site testing data while the cases from the other four PET sites and the other three SPECT sites were used for model development.** (A): hyperparameter selection pipeline, (B): model training and selection pipeline, (C): model testing pipeline. PET – positron emission tomography, SPECT – single-photon emission computed tomography, BWH – Brigham and Women's Hospital, ITMT – Intermountain Healthcare, CSMC – Cedars-Sinai Medical Center, MYAO – Mayo Clinic, NAPLES – University of Naples Federico II, Yale – Yale University, CALGARY – University of Calgary, COLUMBIA – Columbia University, OTTAWA – University of Ottawa, AUC – area under receiver operating characteristic curve, XGBoost – eXtreme Gradient Boosting

(A) Correlation coefficient among hepatic measures

(B) Correlation coefficient between hepatic measures and existing risk variables

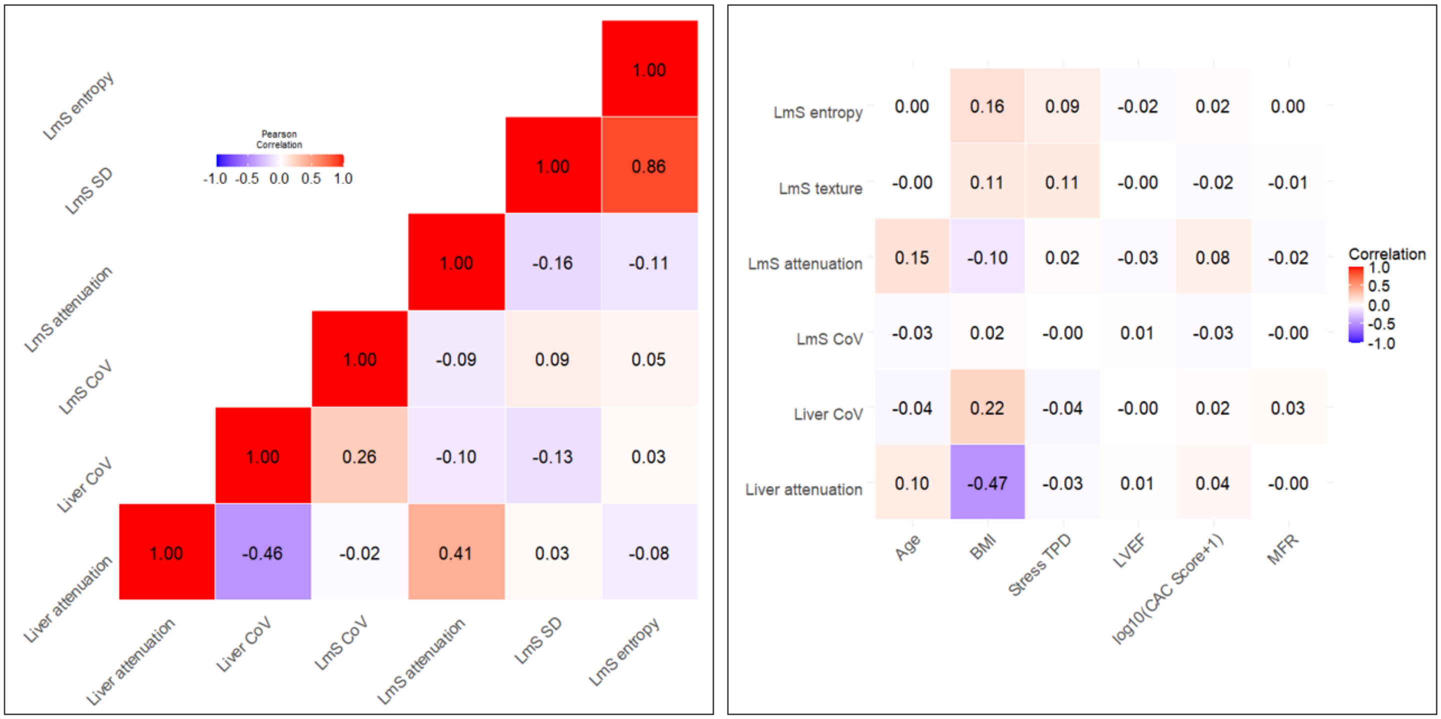


**Supplementary Figure 2. Correlation plots of variables with VIF < 5.** (A) among hepatic measures; (B) between hepatic measures and clinical/imaging variables. Correlation with MFR was only calculated for PET patients. Correlation coefficient for MFR was only calculated within PET patients. VIF – variance inflation factor, LmS – difference of liver minus spleen, SD – standard deviation, CoV – coefficient of variation, BMI – body mass index, TPD – total perfusion deficit, LVEF – left ventricular ejection fraction, CAC – coronary artery calcium, MFR – myocardial flow reserve.

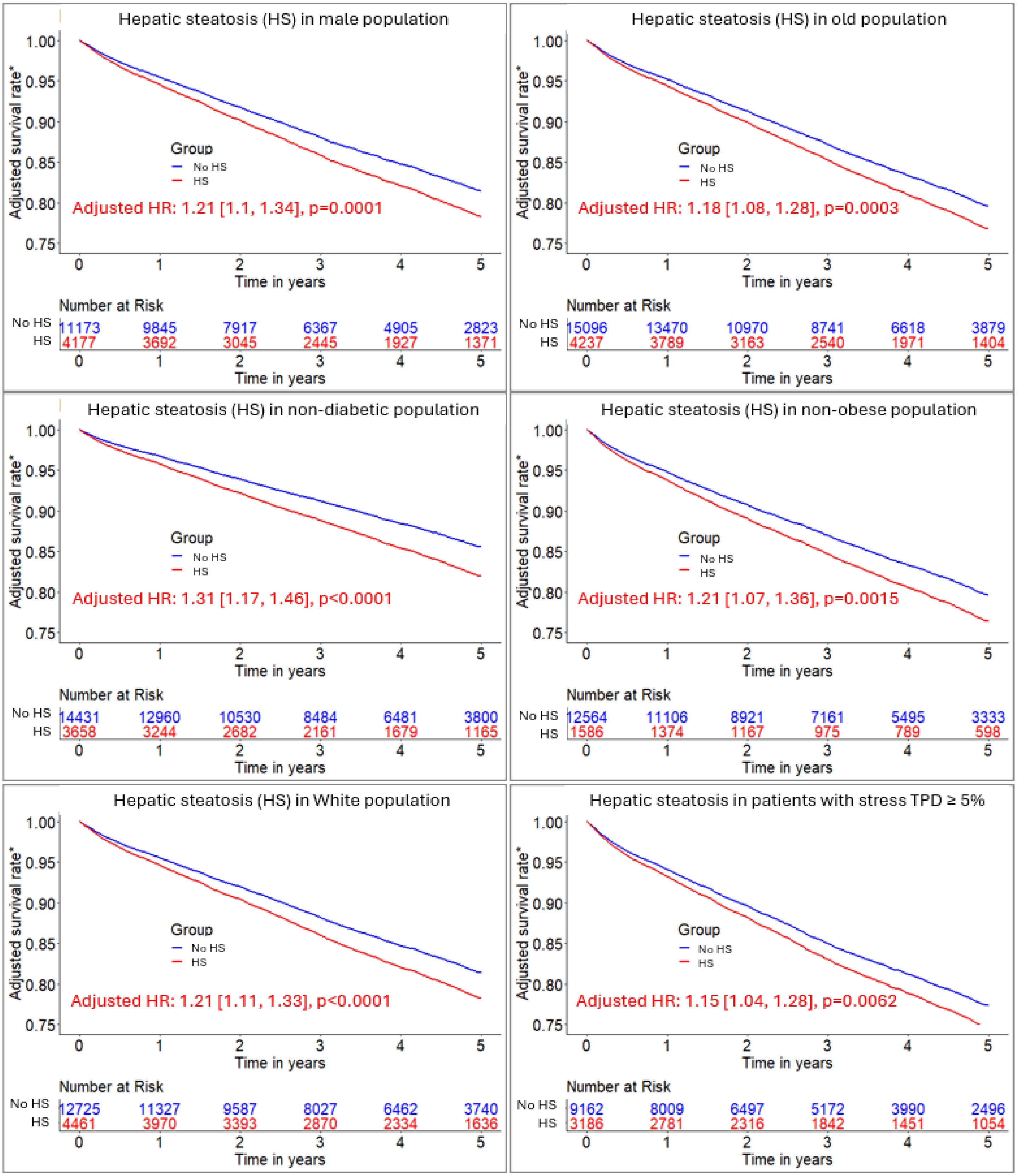


**Supplementary Figure 3. Adjusted Kaplan-Meier curves of hepatic steatosis in male, old, non-obese, White, non-diabetic, and abnormal-perfusion populations, respectively from a cohort of 27039 patients.** Patients without hepatic steatosis measures were reference groups. Clinical and perfusion factors: sex, age, body mass index, hypertension, diabetes mellitus, dyslipidemia, family history of coronary artery disease, smoking, stress total perfusion deficit, left ventricle ejection fraction, and deep-learning-coronary artery calcium score. Patients were considered old if they had age ≥ 60 years, and non-obese if they had body mass index ≥ 30 kg/m². *Adjusted for clinical, perfusion factors and all the other hepatic quantifications. HR of age/BMI/log10(CAC score+1) was per unit increase. Female/LVEF ≥ 50%/Stress TPD<5%/low liver CoV/low LmS CoV/low LmS entropy/low LmS SD was reference group. For diabetes mellitus, dyslipidemia, family history of CAD, hypertension, and smoking, patients without them were the reference groups. Abnormal perfusion was defined as stress TPD ≥ 5%. BMI – body mass index, CAD – coronary artery disease, TPD – total perfusion deficit, LVEF – left ventricular ejection fraction, CAC – coronary artery calcium

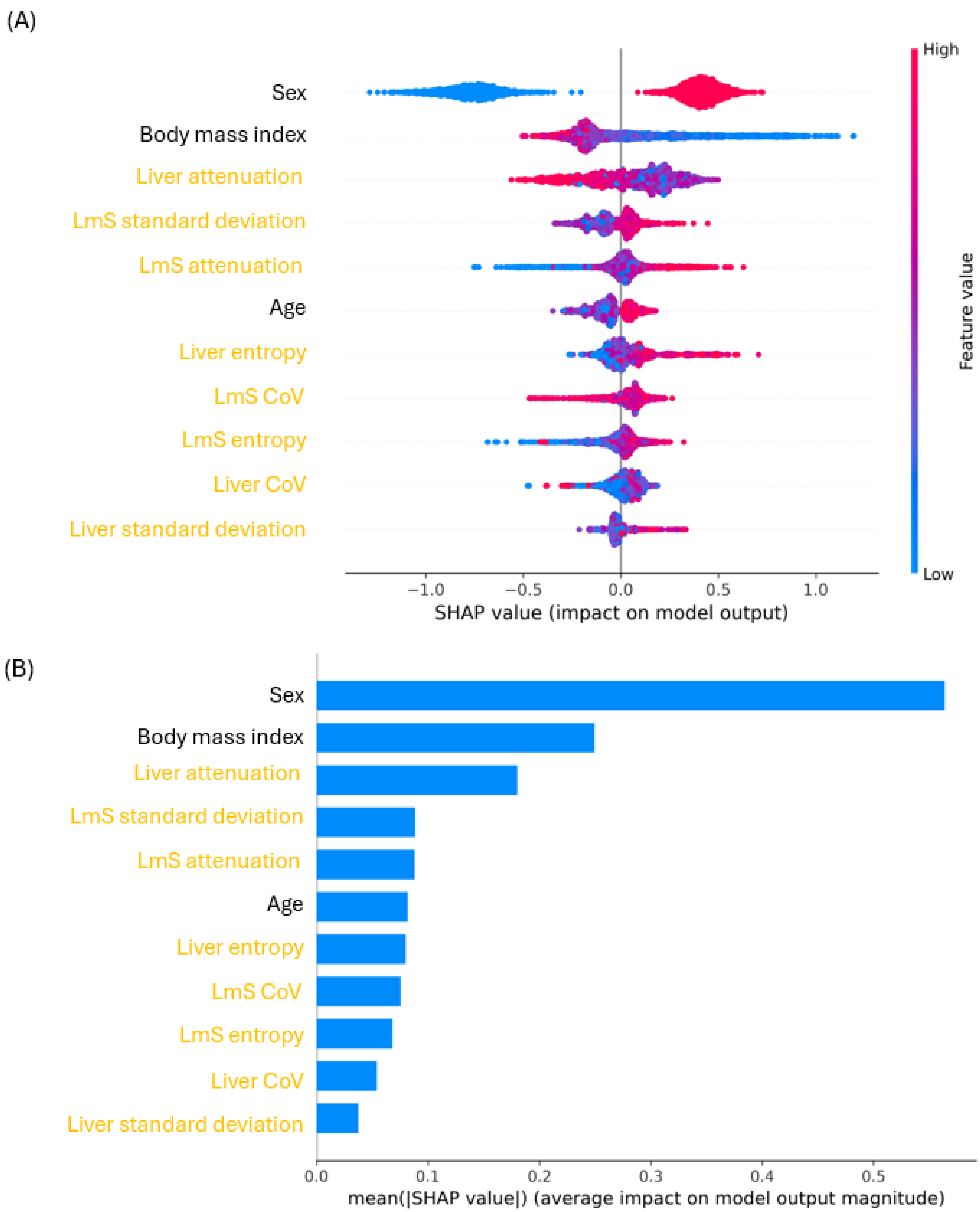


**Supplementary Figure 4. Contribution of individual hepatic measure to liver risk index (LIRI).** (A): scatter plot of SHAP value; (B): normalized mean absolute mean plot of SHAP value. Yellow indicates hepatic measures. Hepatic steatosis (HS) was determined according to the imaging criterion: a patient had HS if liver attenuation < 40 HU or liver minus spleen (LmS) attenuation < -10. LVEF

– left ventricular ejection fraction, CAC – coronary artery calcium, TPD – total perfusion deficit, CoV – coefficient of variation, CAD – coronary artery disease.
